# Genome-wide Association Identifies Novel Etiological Insights Associated with Parkinson’s Disease in African and African Admixed Populations

**DOI:** 10.1101/2023.05.05.23289529

**Authors:** Mie Rizig, Sara Bandres-Ciga, Mary B Makarious, Oluwadamilola Ojo, Peter Wild Crea, Oladunni Abiodun, Kristin S Levine, Sani Abubakar, Charles Achoru, Dan Vitale, Olaleye Adeniji, Osigwe Agabi, Mathew J Koretsky, Uchechi Agulanna, Deborah A. Hall, Rufus Akinyemi, Tao Xie, Mohammed Ali, Ejaz A. Shamim, Ifeyinwa Ani-Osheku, Mahesh Padmanaban, Owotemu Arigbodi, David G Standaert, Abiodun Bello, Marissa Dean, Cyril Erameh, Inas Elsayed, Temitope Farombi, Olaitan Okunoye, Michael Fawale, Kimberley J Billingsley, Frank Imarhiagbe, Pilar Alvarez Jerez, Emmanuel Iwuozo, Breeana Baker, Morenikeji Komolafe, Laksh Malik, Paul Nwani, Kensuke Daida, Ernest Nwazor, Abigail Miano-Burkhardt, Yakub Nyandaiti, Zih-Hua Fang, Yahaya Obiabo, Jillian H. Kluss, Olanike Odeniyi, Dena Hernandez, Francis Odiase, Nahid Tayebi, Francis Ojini, Ellen Sidranksy, Gerald Onwuegbuzie, Andrea M. D’Souza, Godwin Osaigbovo, Bahafta Berhe, Nosakhare Osemwegie, Xylena Reed, Olajumoke Oshinaike, Hampton Leonard, Folajimi Otubogun, Chelsea X Alvarado, Shyngle Oyakhire, Simon Ozomma, Sarah Samuel, Funmilola Taiwo, Kolawole Wahab, Yusuf Zubair, Hirotaka Iwaki, Jonggeol Jeffrey Kim, Huw R Morris, John Hardy, Mike Nalls, Karl Heilbron, Lucy Norcliffe-Kaufmann, Nigeria Parkinson Disease Research Network, International Parkinson’s Disease Genomics Consortium - Africa (IPDGC Africa), Black and African American Connections to Parkinson’s Disease (BLAAC PD) Study Group, the 23andMe Research Team, Cornelis Blauwendraat, Henry Houlden, Andrew Singleton, Njideka Okubadejo, Global Parkinson’s Genetics Program

**Affiliations:** Department of Neuromuscular Diseases, UCL Queen Square Institute of Neurology, London WC1N 3BG, UK; UCL Movement Disorders Centre, University College London, London, WC1N 3BG, UK; Center for Alzheimer’s and Related Dementias (CARD), National Institute on Aging and National Institute of Neurological Disorders and Stroke, National Institutes of Health, Bethesda, MD, USA, 20814; Laboratory of Neurogenetics, National Institute on Aging, National Institutes of Health, Bethesda, MD, USA; College of Medicine, University of Lagos, Idi Araba, Lagos State, Nigeria; General Hospital, Isolo, Lagos State, Nigeria; Data Tecnica International, Washington, DC, USA; Ahmadu Bello University, Zaria, Kaduna State, Nigeria; Jos University Teaching Hospital, Jos, Plateau State, Nigeria; Federal Medical Centre, Abeokuta, Ogun State, Nigeria; Lagos University Teaching Hospital, Idi Araba, Lagos State, Nigeria; Department of Neurological Sciences, Rush University Medical Center, Chicago, IL, USA; Neuroscience and Ageing Research Unit, Institute for Advanced Medical Research and Training, College of Medicine, University of Ibadan, Ibadan, Oyo State, Nigeria; Department of Neurology, University of Chicago Medicine, Chicago, Illinois, USA; Federal Teaching Hospital Gombe, Gombe State, Nigeria; Human Motor Control Section, National Institute of Neurological Disorders and Stroke, National Institutes of Health, Bethesda, Maryland, USA; Kaiser Permanente Mid-Atlantic States, Largo, Maryland, USA; MidAtlantic Permanente Research Institute, Rockville, Maryland, USA; Asokoro District Hospital, Asokoro, Abuja, Nigeria; Delta State University, Abraka, Delta State, Nigeria; Department of Neurology, University of Alabama at Birmingham, Birmingham, AL, USA; University of Ilorin Teaching Hospital, Ilorin, Kwara State, Nigeria; Irrua Specialist Teaching Hospital, Irrua, Edo State, Nigeria; Faculty of Pharmacy, University of Gezira, Wadmadani, 20, Sudan; University College Hospital, Ibadan, Oyo State, Nigeria; Obafemi Awolowo University, Ile-Ife, Osun State, Nigeria; University of Benin, Benin City, Edo State, Nigeria; Benue State University, Makurdi, Benue State, Nigeria; Nnamdi Azikiwe University Teaching Hospital, Nnewi, Anambra State, Nigeria; Rivers State University Teaching Hospital, Port Harcourt, Rivers State, Nigeria; University of Maiduguri Teaching Hospital, Maiduguri, Borno State, Nigeria; German Center for Neurodegenerative Diseases (DZNE), Tuebingen, Germany; Federal University of Health Sciences, Otukpo, Benue State, Nigeria; General Hospital, Lagos Island, Lagos State, Nigeria; Medical Genetics Branch, National Human Genome Research Institute, National Institutes of Health, Bethesda, Maryland, USA; University of Abuja, Abuja, Federal Capital Territory, Nigeria; University of Port Harcourt, Port Harcourt, Rivers State, Nigeria; Lagos State University College of Medicine, Ikeja, Lagos State, Nigeria; Federal Medical Center, Ebute Metta, Lagos State, Nigeria; National Hospital, Abuja, Federal Capital Territory, Nigeria; University of Calabar Teaching Hospital, Calabar, Cross River State, Nigeria; University of Ilorin, Ilorin, Kwara State, Nigeria; 23andMe, Inc., Sunnyvale, CA, USA

**Author notes:** Correspondence to: Njideka Okubadejo, MD | Professor & Consultant Neurologist College of Medicine, University of Lagos & Lagos University Teaching Hospital, Idi Araba, Lagos State, Nigeria Andrew Singleton, PhD | Director, Center for Alzheimer’s and Related Dementias National Institute on Aging and National Institute of Neurological Disorders and Stroke, National Institutes of Health, Bethesda, MD, USA. joint first (Mie Rizig, Sara Bandres-Ciga, Mary B Makarious). joint last (Andrew Singleton, Njideka Okubadejo). Funding: Data used in the preparation of this article were obtained from Global Parkinson’s Genetics Program (GP2). GP2 is funded by the Aligning Science Across Parkinson’s (ASAP) initiative and implemented by The Michael J. Fox Foundation for Parkinson’s Research (https://gp2.org). For a complete list of GP2 members see https://gp2.org. Additional funding was provided by The Michael J. Fox Foundation for Parkinson’s Research through grant MJFF-009421/17483.

**Keywords:** genetics, Parkinson’s disease, genome-wide association study, African, African Admixed, *GBA1*, expression quantitative trait locus, therapeutic interventions.

## Abstract

**Background:** Understanding the genetic mechanisms underlying diseases in ancestrally diverse populations is a critical step towards the realization of the global application of precision medicine. The African and African admixed populations enable mapping of complex traits given their greater levels of genetic diversity, extensive population substructure, and distinct linkage disequilibrium patterns.

**Methods:** Here we perform a comprehensive genome-wide assessment of Parkinson’s disease (PD) in 197,918 individuals (1,488 cases; 196,430 controls) of African and African admixed ancestry, characterizing population-specific risk, differential haplotype structure and admixture, coding and structural genetic variation and polygenic risk profiling.

**Findings:** We identified a novel common risk factor for PD and age at onset at the *GBA1* locus (risk, rs3115534-G; OR=1.58, 95% CI = 1.37 - 1.80, P=2.397E-14; age at onset, BETA =-2.004, SE =0.57, P = 0.0005), that was found to be rare in non-African/African admixed populations. Downstream short- and long-read whole genome sequencing analyses did not reveal any coding or structural variant underlying the GWAS signal. However, we identified that this signal mediates PD risk via expression quantitative trait locus (eQTL) mechanisms. While previously identified *GBA1* associated disease risk variants are coding mutations, here we suggest a novel functional mechanism consistent with a trend in decreasing glucocerebrosidase activity levels. Given the high population frequency of the underlying signal and the phenotypic characteristics of the homozygous carriers, we hypothesize that this variant may not cause Gaucher disease. Additionally, the prevalence of Gaucher’s disease in Africa is low.

**Interpretation:** The present study identifies a novel African-ancestry genetic risk factor in *GBA1* as a major mechanistic basis of PD in the African and African admixed populations. This striking result contrasts to previous work in Northern European populations, both in terms of mechanism and attributable risk. This finding highlights the importance of understanding population-specific genetic risk in complex diseases, a particularly crucial point as the field moves toward precision medicine in PD clinical trials and while recognizing the need for equitable inclusion of ancestrally diverse groups in such trials. Given the distinctive genetics of these underrepresented populations, their inclusion represents a valuable step towards insights into novel genetic determinants underlying PD etiology. This opens new avenues towards RNA-based and other therapeutic strategies aimed at reducing lifetime risk.

**Research in Context:** 

**Evidence Before this Study:** Our current understanding of Parkinson’s disease (PD) is disproportionately based on studying populations of European ancestry, leading to a significant gap in our knowledge about the genetics, clinical characteristics, and pathophysiology in underrepresented populations. This is particularly notable in individuals of African and African admixed ancestries. Over the last two decades, we have witnessed a revolution in the research area of complex genetic diseases. In the PD field, large-scale genome-wide association studies in the European, Asian, and Latin American populations have identified multiple risk loci associated with disease. These include 78 loci and 90 independent signals associated with PD risk in the European population, nine replicated loci and two novel population-specific signals in the Asian population, and a total of 11 novel loci recently nominated through multi-ancestry GWAS efforts.

Nevertheless, the African and African admixed populations remain completely unexplored in the context of PD genetics.

**Added Value of this Study:** To address the lack of diversity in our research field, this study aimed to conduct the first genome-wide assessment of PD genetics in the African and African admixed populations. Here, we identified a genetic risk factor linked to PD etiology, dissected African-specific differences in risk and age at onset, characterized known genetic risk factors, and highlighted the utility of the African and African admixed risk haplotype substructure for future fine-mapping efforts. We identified a novel disease mechanism via expression changes consistent with decreased *GBA1* activity levels. Future large scale single cell expression studies should investigate the neuronal populations in which expression differences are most prominent. This novel mechanism may hold promise for future efficient RNA-based therapeutic strategies such as antisense oligonucleotides or short interfering RNAs aimed at preventing and decreasing disease risk. We envisage that these data generated under the umbrella of the Global Parkinson’s Genetics Program (GP2) will shed light on the molecular mechanisms involved in the disease process and might pave the way for future clinical trials and therapeutic interventions. This work represents a valuable resource in an underserved population, supporting pioneering research within GP2 and beyond. Deciphering causal and genetic risk factors in all these ancestries will help determine whether interventions, potential targets for disease modifying treatment, and prevention strategies that are being studied in the European populations are relevant to the African and African admixed populations.

**Implications of all the Available Evidence:** We nominate a novel signal impacting *GBA1* as the major genetic risk factor for PD in the African and African admixed populations. The present study could inform future *GBA1* clinical trials, improving patient stratification. In this regard, genetic testing can help to design trials likely to provide meaningful and actionable answers. It is our hope that these findings may ultimately have clinical utility for this underrepresented population.

## Introduction

Parkinson’s disease (PD) is a complex, heterogeneous neurodegenerative disorder that manifests with progressive motor and non-motor features, including resting tremor, bradykinesia, mood disorders, olfactory dysfunction, and cognitive impairment. Globally, about 6.1 million people had PD in 2016^1^, and as a result of an aging world population and increased longevity, this figure is expected to rise to 17.5 million by 2040^2^ as a result of an aging world population and increased longevity.

Genome-wide association studies (GWAS) have been instrumental for identifying common variants associated with complex diseases like PD, unraveling the genetics and heritability of PD in European populations^3^. The largest published GWAS meta-analysis of PD risk to date was performed on individuals of European ancestry and identified 90 independent genome-wide significant risk signals that explain 16-22% of the heritable risk of PD^4, 5^. However, very little is known about the genetics of PD in non-European populations. The largest PD GWAS meta-analysis in the East Asian population recently identified two population-specific signals,^6^ and the first PD GWAS in Latin Americans has suggested two potential novel loci that warrant further study^7^. The first multi-ancestry PD GWAS meta-analysis has nominated 11 novel loci, providing a foundation for future efforts aimed at fine-mapping novel genetic regions linked to PD^8^. GWAS are powerful tools in the creation of better prediction models and broadening our biological knowledge of specific diseases^9^.

Nearly one-third of the genetic heritability of PD can be explained by polygenic risk scores (PRSs) according to the most recent genetic studies conducted in Europeans. However, the heritability explained by PRSs is totally unknown in under-researched and underserved populations, as is the total heritability^10, 11^. There has been considerable ethnic variability in the distribution of monogenic causes and genetic risk variants documented across populations. For instance, the relatively common LRRK2 p.G2019S mutation remains unreported in some sub-Saharan African populations, despite being most commonly associated with familial and sporadic PD in Zambia and Northern Africa^12–16^.

African and African admixed populations offer unique opportunities for studying the genetics of both monogenic and complex diseases because they contain the largest portion of the within-population genetic variability in the world, shorter linkage disequilibrium (LD) blocks, and abundant alleles that are private to these populations^17–19^. In addition to promoting scientific equity to address health disparities, diverse representation provides a platform for replication studies to explore the strength and relevance of findings reported from other populations. Additionally it has the potential to facilitate the identification of novel or unique loci and investigate genotype-phenotype correlations that can further expand our understanding of pathological and pathogenetic disease mechanisms in PD^17, 20^.

This study provides the first GWAS-based insights into the genetics of PD in the African and African admixed populations **(Figure 1)**. Here we performed a comprehensive genome-wide assessment of PD risk and age at onset, characterizing population specific cumulative risk profiling, haplotype structure, and genetic admixture. Leveraging this unique population genetic structure, our analyses identified a novel association signal in *GBA1, t*he gene encoding the lysosomal enzyme glucocerebrosidase (GCase). This led to the investigation of a novel disease mechanism of expression changes consistent with decreased glucocerebrosidase activity levels relating to increased risk. Finally, we compare our findings in the context of other global populations. We envisage that these data generated under the umbrella of the Global Parkinson’s Genetics Program (GP2) will shed light on the molecular mechanisms involved in the disease process and might pave the way for future RNA-based therapeutic strategies aimed at reducing lifetime risk.

**Figure 1.**
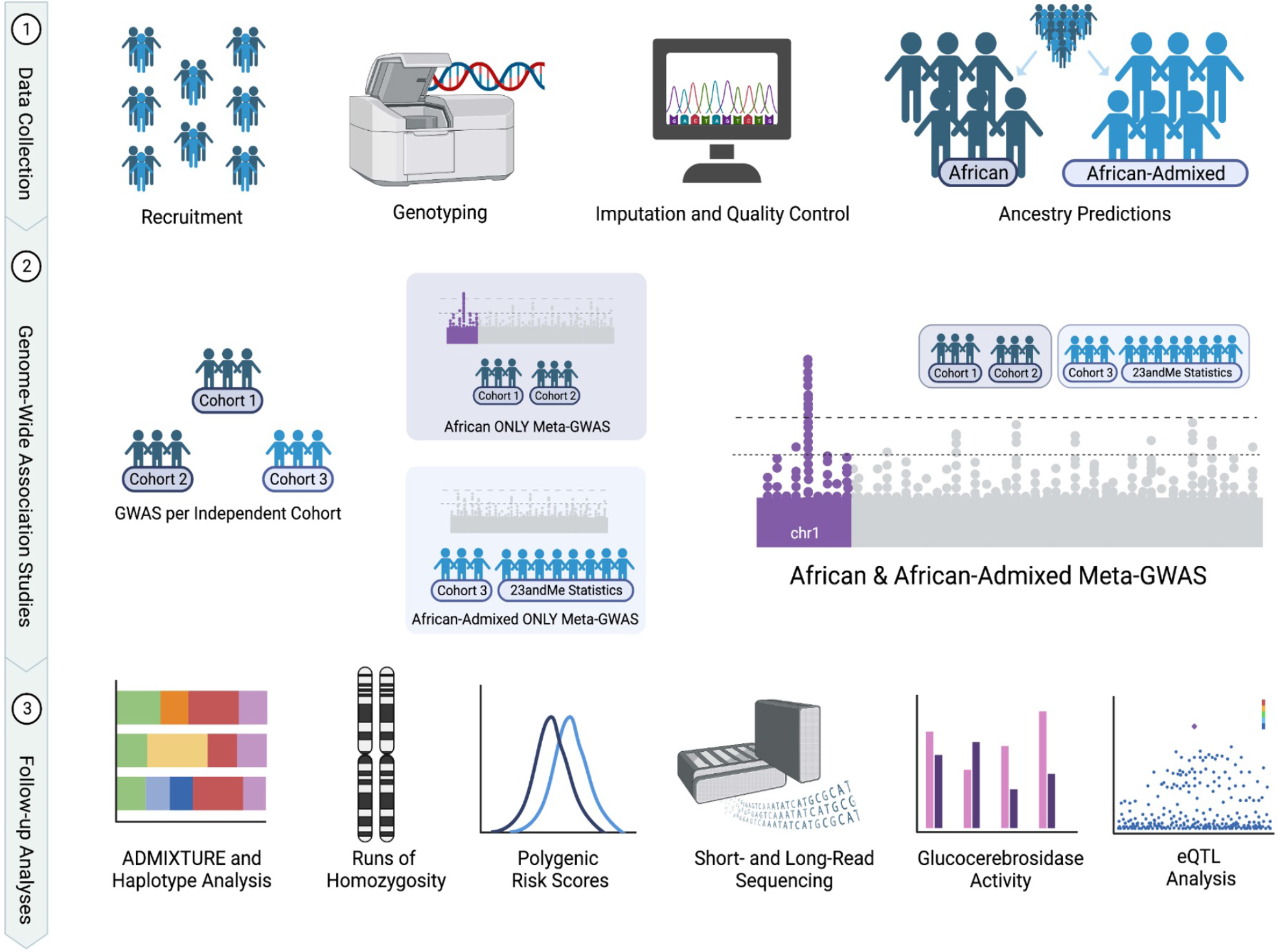
Analysis Workflow Schematic

## Methods

### Study participants

The demographic and clinical characteristics of the cohorts under study are provided in **Table 1.** Three sources of data were included in this study: Individual level data from the International Parkinson’s Disease Genomics Consortium - Africa *(IPDGCAN)* and the Global Parkinson’s Disease Genetics Program *(GP2),* and GWAS summary statistics from *23andMe, Inc*.

**Table 1.**
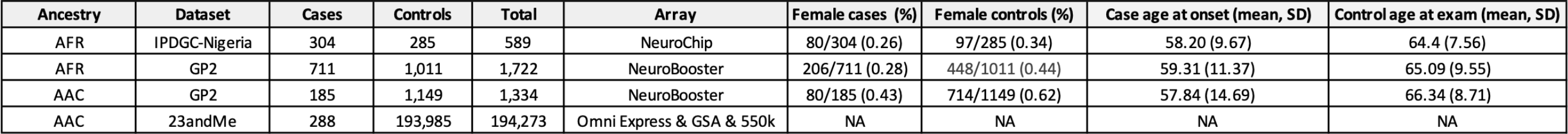
Demographic and clinical characteristics of the cohorts under study

For the *IPDGCAN* and the *GP2* cohorts, the diagnosis of PD was based on fulfillment of the United Kingdom PD Society Brain Bank criteria (excluding the requirement for not more than one affected relative)^21^. The respective ethical committees for medical research approved involvement in genetic studies, and participants gave informed written consent. All participants underwent a neurological examination conducted by a study neurologist to document clinical and neurological status. Controls were generally assessed to detect overall signs of neurological condition and samples presenting any clinical signs of neurodegenerative diseases were excluded from the control series.

Summary statistics for individuals with or without PD were provided through a collaborative agreement with 23andMe, Inc. Participants provided informed consent and volunteered to participate in the research online, under a protocol approved by the external AAHRPP-accredited IRB, Ethical & Independent (E&I) Review Services. As of 2022, E&I Review Services is part of Salus IRB (https://www.versiticlinicaltrials.org/salusirb). PD diagnosis was self-reported in this instance. A previous PD GWAS meta-analysis found a strong genetic correlation between 23andMe GWAS data using self-reported cases and non-23andMe GWAS data with PD cases ascertained by clinicians (genetic correlation from LDSC (rG) = 0.85, SE = 0.06)^4^. Age distributions of cohorts under study are illustrated in **Supplementary Figure 1.**

### Genotype data generation, quality control, ancestry predictions, and imputation

*The IPDGCAN* and *GP2* samples were genotyped using two different genotyping platforms **(Table 1).** The NeuroBooster array (v.1.0, Illumina, San Diego, CA) contains a backbone of 1,914,935 variants densely covering ancestry informative markers, markers for determination of identity by descent, and X-chromosome SNPs for sex determination. In addition, it contains 96,517 customized variants. Samples collected as part of the *GP2* initiative were genotyped on this array. Samples collected as part of the *IPDGCAN* initiative **(Table 1)** were genotyped using two different platforms; the Neurochip array, containing a backbone of 306,670 variants and customized content comprising 179,467 variants^22^, and the previously described NeuroBooster array.

Raw genotype data was passed through a custom ancestry prediction and pruning machine learning method as a part of the GenoTools pipeline (https://github.com/GP2code/GenoTools), as described elsewhere^23^. All samples underwent similar standardized quality control (QC) as follows: Samples were excluded from the analysis if: call rate was <95%, genetically determined sex did not match that from clinical data, or excess heterozygosity was detected (|F| statistics > 0.25).

Samples were subset by ancestry estimates (see **Supplementary Materials** for details). In brief, ancestry was defined using reference panels from the 1000 Genomes Project^24^, Human Genome Diversity Project^25^, and an Ashkenazi Jewish population dataset^26^. In total, 39,302 reference panel SNPs were genotyped on the NeuroBooster array and 24,404 reference panel SNPs were also genotyped on the NeuroChip array (see **Supplementary Materials** for details). Ancestry estimates were carried out using a uniform protocol across all samples.

Next, we removed those samples that were IBD for > 12.5% of the genome (approximately related at a first cousins level or closer). Once preliminary sample-level QC was completed, SNPs with Hardy-Weinberg Equilibrium (HWE) P value <1E-4 in control samples were removed. Next, variants were pruned for missingness by case-control status at P≤1E-4 to remove variants with non-random missingness. Finally, variants were pruned for non-random missingness by haplotype at P≤1E-4.

For the *GP2* data, variants were further filtered by minor allele frequency (MAF) < 0.005 and HWE P < 1E-5 prior to being submitted to the TOPMed Imputation server. The TOPMed reference panel version r^2^ contains information from 97,256 reference samples and more than 300 million genetic variants across the 22 autosomes and the X chromosome. As of October 2022, the TOPMed panel consists of about 180,000 participants of which 29% are of African ancestry, 19% of Latin American ancestry, 8% of Asian ancestry, and 40% of European ancestry. More information about the TOPMed Study^27^, Imputation Server^28^, and Minimac Imputation^29^ can be found at https://imputation.biodatacatalyst.nhlbi.nih.gov.

The imputed files were then pruned applying a minor allele count (MAC) threshold of 10 and an imputation Rsq of 0.3. For additional information regarding *GP2* ancestry prediction as well as 23andMe data generation and processing, please see the **Supplementary Materials.**

### Estimation of PD risk, age at onset and admixture

To estimate risk associated with PD, imputed dosages (meaning genotype probabilities for a variant to be A/A, A/B, or B/B from 0 to 2 that account for some uncertainty) were analyzed using a logistic regression model adjusted for sex, age, and the first ten PCs as covariates. Age at onset (AAO) was used for cases and age at recruitment was used for controls. In instances where AAO was not available for cases, age at recruitment was used instead (less than 6% of individuals). For individuals who had no age information provided, average age was imputed (less than 5% and 2% of cases and controls, respectively). Summary statistics were generated using PLINK 1.9 and 2.0^34^, and filtered for inclusion after meeting a minimum imputation quality of 0.30 and MAF > 5 %. A binomial generalized linear model (GLM) was employed to assess the predictive ability of polygenic risk score (PRS) between cases and controls, taking into account demographic variables such as age, sex, and the first ten PCs as covariates. To explore the influence of genetic variation on the AAO of PD cases, a linear regression model adjusted for the same covariates was performed. Additionally, we conducted linear regression analyses to explore how potential GWAS signals would correlate with admixture levels. All the analyses were performed on Terra (https://terra.bio/).

GWAS was conducted on African and African admixed ancestries independently and then meta-analyzed. We utilized fixed-effects meta-analyses as implemented in METAL^35^ to leverage summary statistics across all sources. Pairwise LD values were calculated using 1000 genomes African population data through LD link (https://ldlink.nci.nih.gov/?tab=home).

### Haplotype and fine-mapping analyses

Haplotype size was compared using individual level data across African, African admixed, and European PD cases. After standardizing the three datasets with the same genotyped SNPs passing identical QC steps, we determined the size of the haplotype blocks using default parameters in PLINK 1.9. This analysis estimates haplotype blocks by Haploview’s interpretation of the block definition. By default, only pairs of variants within 200 kilobases (kb) of each other were considered. Two variants are considered by this procedure to be in strong LD if the lower bound of the 90% D-prime confidence interval (CI) was >0.70, and the upper bound of the CI was at least 0.98.

In an attempt to prioritize putative causal variants within the identified *GBA1* risk haplotype, we performed fine-mapping analyses across the LD block where the genome-wide signal was located by using the “Approximate Bayes Factor fine mapping under a single causal variant assumption” method provided by the R package coloc (https://CRAN.R-project.org/package=coloc). This analysis assesses the posterior probability of each SNP being the causal variant within a locus. We derived posterior probabilities (PP) for this region using the default prior probability of 1E-4 under the assumption of a single causative variant per locus.

### Short-read Whole Genome Sequencing

To further dissect the novel identified GWAS signal, we performed whole-genome sequencing (WGS) analyses in 206 individuals (141 cases and 65 controls) of which 39 individuals were *GBA1* rs3115534-GG carriers, 69 were rs3115534-GT and 98 were rs3115534-TT carriers. Short-read WGS DNA sequencing was performed by Psomagen (detailed in Supplementary methods). We used the functional equivalence pipeline^36^ implemented at the Broad Institute to produce alignments and small variant calls against the GRCh38DH reference genome. For sample-level WGS quality control, we followed the quality metrics defined by the Accelerating Medicines Partnership Parkinson’s Disease initiative (AMP-PD;

https://amp-pd.org^37^. To produce a set of joint-genotyped variants for all the samples that passed quality control, we ran the Broad Institute’s joint discovery pipeline and retained only the high-quality variants flagged as “PASS” after variant quality score recalibration, with a call rate > 0.95, genotype quality >20, read depth >5, and heterozygous allele balance between 0.25 and 0.75 as described previously ^38^. Additionally, we called *GBA1* variants using Gauchian v1.0.2^39^ and genotyped known neurological repeat expansions using STRipy v2.2^40^. All the pipelines and scripts used are available via GitHub (https://github.com/GP2code). Data passing quality control metrics were annotated using ANNOVAR^33^. A comprehensive assessment of known and potential novel pathogenic variants driving the *GBA1* signal was performed. CRAM files were visualized using the Integrative Genomics Viewer (IGV) web browser^41^.

The Gauchian algorithm^39^ was then applied to nominate potential structural variants driving the *GBA1* signal. Briefly, this algorithm is a targeted variant caller for the *GBA1* gene based on WGS BAM files. Gauchian aims to solve the problems caused by the high sequence similarity with the pseudogene paralog *GBAP1*. This algorithm has been reported to be able to detect variants in the exons 9-11 homology region, such as large deletions or duplications between *GBA1* and *GBAP1*, and *GBAP1*-like variants in *GBA1*, including p.A495P, p.L483P, p.D448H, c.1263del, RecNciI, RecTL and c.1263del+RecTL.

### Long-read Whole Genome Sequencing

Oxford Nanopore Technologies (ONT) long-read whole-genome sequencing data was generated for five *GBA1* rs3115534-GG carriers, two heterozygotes and six *GBA1* rs3115534-TT carriers. High molecular weight DNA was extracted from either frozen blood samples or cell-lines. For the blood samples DNA was extracted from 1ml per sample using the Kingfisher APEX instrument with the Nanobind CBB Big DNA kit (HBK-CBB-001). For the frozen cell-pellets DNA was extracted manually with the Nanobind CBB Big DNA kit (HBK-CBB-001) using the following protocol^42^(https://dx.doi.org/10.17504/protocols.io.q26g74169gwz/v1).

The DNA then went through a size selection step using the Circulomics Short Read Eliminator Kit (SS-100-101-01) to remove fragments up to 25kb. Finally a library was prepared with the SQK-LSK 110 Ligation Sequencing Kit from ONT and each library was loaded onto a separate PromethION R9.4.1 flow cell following ONT standard operating procedures and ran for a total of 72 hours on a PromethION device.

Fast5 files containing raw signal data were obtained from sequencing performed using minKNOW v22.10.7 (ONT). All fast5 files were used to conduct super accuracy basecalling on each sample with Guppy v6.12. Fastq files that passed quality control filters in the super accuracy base calling step were then mapped to the GRChg38 reference genome using winnowmap v2.03^43^. Structural variants were called with Sniffles2^44^ v2.0.3 using default parameters and the “–tandem-repeats’’ option.

### Glucocerebrosidase activity

Patient-derived lymphoblastoid cell lines (LCLs) were obtained from the Coriell repository (https://www.coriell.org/). LCLs were maintained as directed in suspension with RPMI 1640 (ThermoFisher Scientific, 11875093) containing 2mM Glutamax (ThermoFisher Scientific, 35050061), and 15% FBS (ThermoFisher Scientific, A3160501) at 37°C in 5% CO2. Protein was extracted from LCLs using a citrate-phosphate buffer (0.2 M Na2HPO4, 0.1 M citrate, protease inhibitor, pH 5.8, Millipore Sigma, 11836170001) that was activated with 0.25% Triton X-100. Cells were subjected to a 4-methylumbelliferone (4-MU, Sigma Aldrich, M1381) fluorometric glucocerebrosidase (GCase) activity assay in quadruplicate as previously reported in the literature^45^ with adjusted incubation time of 2.5 hours. A total of 5E6 cells were used per sample with protein concentrations normalized to 0.7 mg/ml via BCA Protein Assay (Thermo Fisher Scientific 23225).

### Polygenic Risk Profiling

Polygenic risk score (PRS) analysis for PD was performed as follows. Briefly, a PRS was calculated incorporating effect estimates from Nalls et al., 2019 summary estimates for the 90 SNPs previously associated with PD risk in European populations^4^. Risk allele dosages were counted, then summed and a genetic risk score was generated across all loci in both African and African admixed data. All SNPs were weighted by their published betas, giving greater weight to alleles with higher risk estimates. PRSs were standardized to have a mean of 0 and standard deviation (SD) of 1. Then, a logistic regression was performed regressing disease status against PRSs. Risk profiling analysis was adjusted for age, sex, and PCs 1-10. We repeated these steps using the African admixed effect estimates from the 23andMe summary statistics for those same 90 SNPs identified by Nalls et al., 2019.

### Runs of Homozygosity

Based on an LD-pruned data set (using previously described parameters), runs of homozygosity (ROHs) were defined using PLINK 1.9 to assess potential over-representation of sharing recessive regions in cases versus controls. Here, we evaluated the largest individual-level dataset (*GP2;* African individuals; **Table 1**), where samples genotyped on the NeuroBooster array included 711 African cases and 1,011 African controls. We explored ROHs containing at least 10 SNPs and a total length ≥1,000 kb, with a rate of scanning windows of at least 0.05 (not containing >1 heterozygous call or 10 missing calls). In order to explore overall homozygosity between cases and controls, three metrics were assessed, including the number of homozygous segments spread across the genome, total kilobase distance spanned by those segments, and average segment size on autosomes only.

## Results

### GWAS reveals a novel genome-wide significant signal associated with PD risk and age at onset

We first performed a GWAS of PD risk in the African population, predominantly consisting of individuals of Nigerian descent which included a total of 997 PD cases and 1,294 controls. Of these individuals, 693 PD cases and 1,009 controls were genotyped on the NeuroBooster array, and 304 PD cases and 285 controls were screened on the NeuroChip array (λ=1.01; **Supplementary Figure 4**). A genome-wide significant SNP at the *GBA1* locus was associated with an increase in PD risk; rs3115534, a variant located in intron 8 of *GBA1* (34 nucleotides upstream of exon 9) was the top hit **(Supplementary Table 1, Supplementary Figure 4;** rs3115534; OR=1.58; 95% CI = 1.35 - 1.84, P=3.44E-09). Contrary to what we would expect when assessing common variation linked to PD risk (MAF > 5%), a high odds ratio was identified for this signal. Our study indicated that each additional risk allele, G, conferred a 1.58 increase in the odds of PD.

In parallel, we performed a GWAS in the African admixed population, leveraging the African-American and Afro-Caribbean datasets available as a part of the GP2 initiative combined with 23andMe African-American summary statistics. The PD African admixed GWAS included a total of 467 PD cases and 195,120 controls (λ=1.01; **Supplementary Figure 5**). No genome-wide significant hits were nominated.

Next, we performed a GWAS meta-analysis of all of the African and African admixed datasets **(Figure 2),** totaling 1,488 cases and 196,430 controls. This revealed that a total of 35 SNPs near the *GBA* gene were significantly associated with PD risk with consistent directionality of effect, the two most distant SNPs being 639,773 base pairs apart from each other. Conditional analyses on the top two SNPs suggested that there is only one causal signal driven by rs3115534 as the leading SNP. Of note, rs3115534-G is much more common in individuals of African or African admixed ancestry relative to other populations; allele frequency = 0.16 according to gnomAD^46^ and allele frequency = 0.21 according to the African 1000 Genomes panel ^24^. The African and African admixed datasets used in this study yielded similar frequencies (African dataset; cohort MAF = 0.25, affected MAF = 0.33, unaffected MAF = 0.19), (African admixed datasets; cohort MAF = 0.14, affected MAF = 0.22, unaffected MAF = 0.13). Within our research cohorts, we found that rs3115534-G was more frequent in Nigerian populations **(Supplementary Table 3).** Linear regression analyses showed that the *GBA1* rs3115534 variant was positively associated with the genome-wide percentage of African ancestry (BETA = -0.001, SE= 0.0005, P= 0.011).

**Figure 2.**
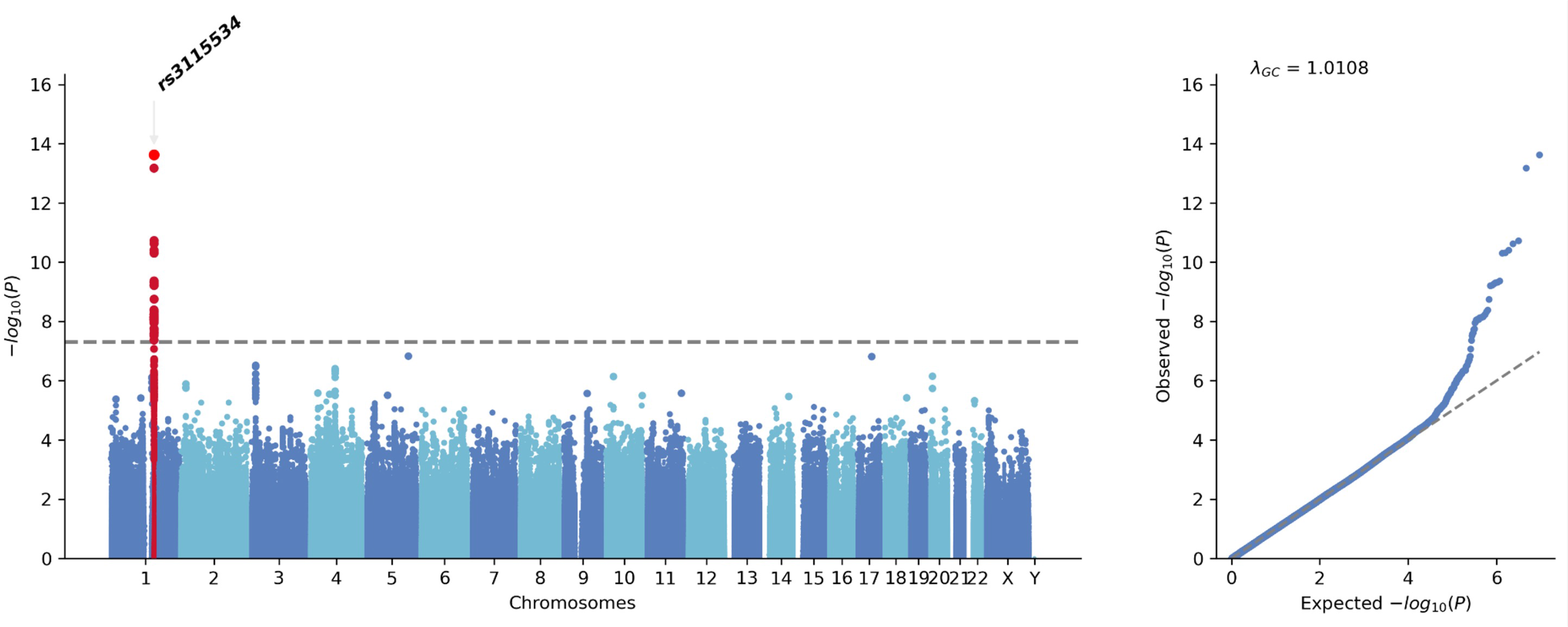
African and African Admixed Caribbean Parkinson’s disease GWAS Meta-analysis

We tested whether the effect of the risk allele was additive by calculating the frequency of homozygotes for the risk allele and heterozygotes in cases versus controls. Notably, our analyses conducted on individual level data from *IPDGCAN* and *GP2* showed that rs3115534-GG was 3.39 times more frequent in African cases (130/1015) than controls (49/1296) and 3.80 times more frequent in African admixed cases (11/185) than controls (18/1149), while rs3115534-GT was 1.17 times more frequent in African cases (398/1015) than controls (435/1296) and 1.38 times more frequent in African admixed cases (61/185) than controls (274/1149). Zygosity analysis of 23andMe data showed that rs3115534-GG was 1.92 times more frequent in African admixed cases (10/288) than controls (3,537/193,985) while rs3115534-GT was 1.27 times more frequent in African admixed cases (85/288) than controls (44,967/193,985). We also analyzed rs3115534 under a dominant model (African ancestry - dominant model: OR = 1.74; 95% CI = 1.40 - 2.15; P = 3.467E-07; African admixed ancestry - dominant model: OR = 1.96; 95% CI = 1.40 - 2.75; P =7.65E-5). Despite the large differences observed in frequencies, effect estimates from the additive model are extremely similar to the dominant model with largely overlapping confidence intervals. This suggests that this variant is additive, and not increasing the risk for PD following a dominant inheritance pattern (African ancestry - additive model: OR =1.75; 95% CI = 1.47 - 2.07, P = 1.40E-10; African admixed ancestry - additive model: OR =1.95; 95% CI =1.47 - 2.60; P =4.12E-6).

As a follow-up analysis, we assessed whether this *GBA1* variant is associated with AAO. Linear regression analyses in 711 African ancestry cases and 185 African admixed ancestry cases showed that *GBA1* rs3115534-G is also an AAO disease modifier (African ancestry: BETA =-2.004, SE = 0.57, P = 0.0005; African-admixed: BETA = -4.15, SE =0.58, P =0.015; Meta-analysis: BETA =-3.06, SE =0.40, P = 0.008) resulting in onset of PD three years earlier per risk allele **(Supplementary Figure 7).** The African-admixed estimates should be taken with caution due to small sample size and low number of GG carriers. No differences in age at PD onset were found between *GBA1* rs3115534-GG and *GBA1* rs3115534-GT carriers (T-test; P = 0.25).

### Genome-wide comparison of the *GBA1* locus across populations suggests an African founder effect

In an attempt to further dissect the novel signal identified in the *GBA1* locus, we next compared effect estimates and directionality of effect leveraging summary statistics from the largest PD GWAS meta-analysis of PD in Europeans^4^, Latin American^7^, and East Asian populations^6^. The rs3115534-G allele is extremely rare in Europeans (allele frequency = 0.0015), East Asians (allele frequency = 0.0005), South Asians (allele frequency =0.0017), and Ashkenazi Jewish populations (allele frequency = 0.0009) according to gnomAD.

When looking at *GP2* European data, the rs3115534 variant was found to be poorly imputed in 13,186 samples (R^2^ = 0.16, MAF = 0.009). In fact, the *GBA1* locus in African and African admixed populations differs substantially from Europeans **(Figure 3; Supplementary Figure 8),** whose association with disease risk is driven by two independent signals, including rs35749011 (*GBA1*-E326K) and rs76763715 (*GBA1*-N370S). These variants are very rare in individuals of African and African admixed ancestry **(Figure 4B).** Similarly, the *GBA1* locus considerably differs from the East Asian population, for which the rs3115534 variant was also not imputed in the largest East Asian GWAS meta-analysis^6^ **(Figure 4C).** These differences are less noticeable when assessing the Amerindian and indigenous populations, which harbor higher levels of African admixture **(Figure 4D)** (Loesch et al. GWAS^7^; rs3115534-G; OR = 1.13, 95% CI =0.41-1.86, P= 0.72; Amerindian and indigenous 23andMe GWAS; rs3115534-G; OR = 1.56, 95% CI = 1.55-1.88, P= 0.01).

**Figure 3.**
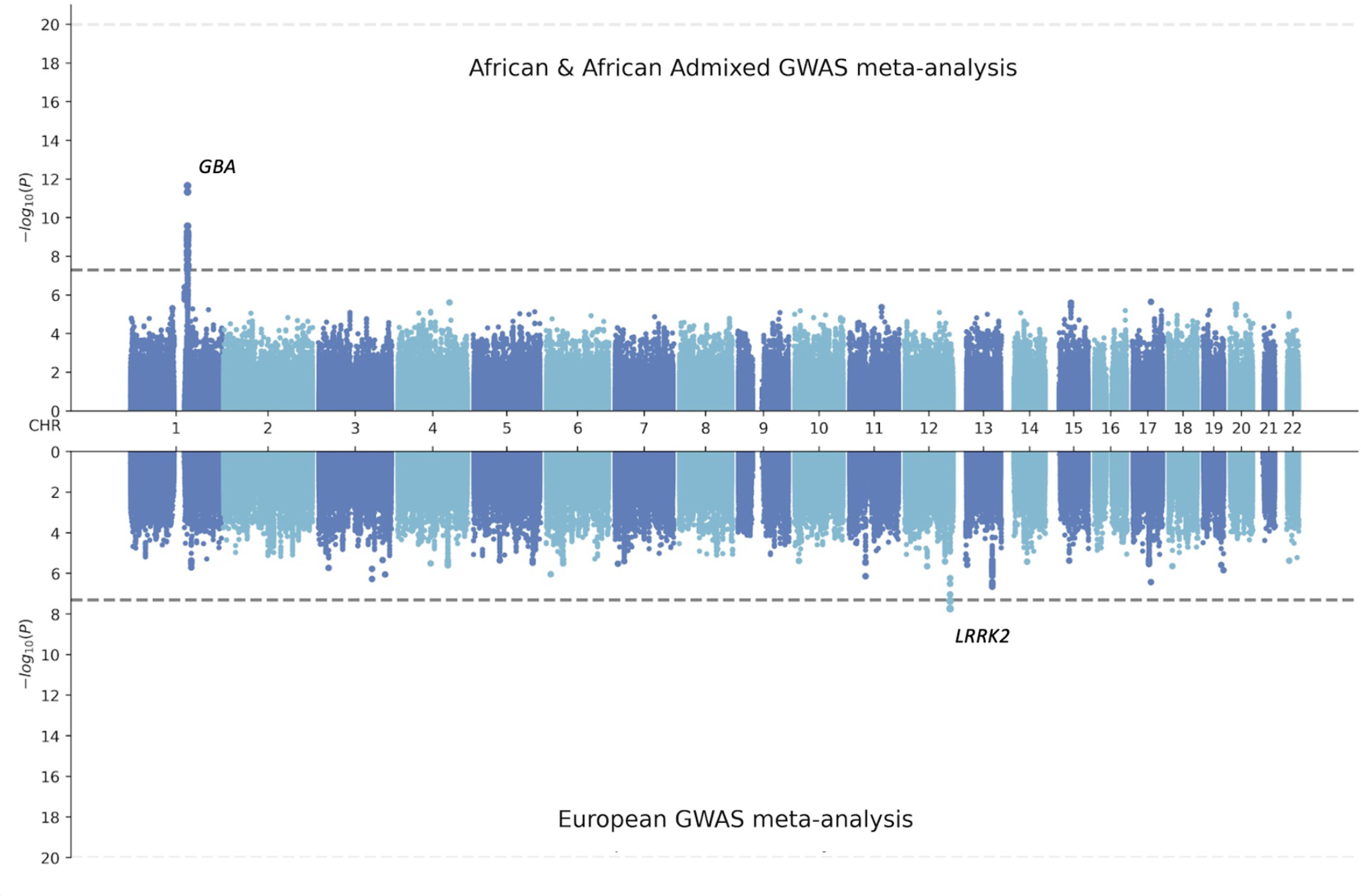
Miami Plot comparing European versus African and African admixed GWAS meta-analysis at similar randomly sampled Parkinson’s disease cases (1,200) and controls (2,445)

**Figure 4.**
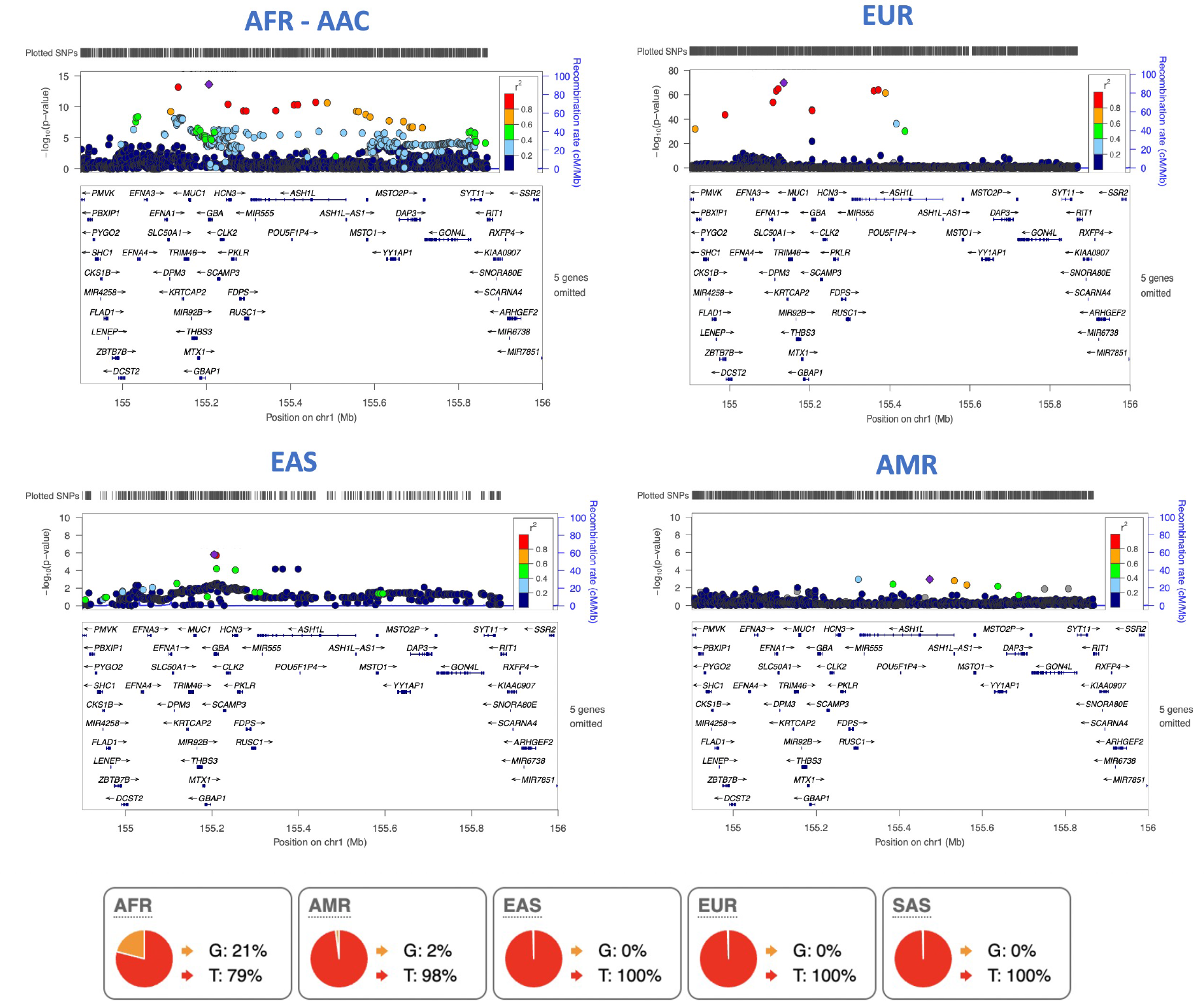
LocusZoom plots of *GBA* in AFR/AAC (A), EUR (B), EAS (C), AMR (D) populations

Furthermore, we assessed the rs3115534-G variant on individual level data from the *GP2* initiative. The variant was not imputed in individuals of European, Ashkenazi Jewish, South Asian, East Asian and Central Asian ancestries, likely due to its low frequency. On the other hand, the rs3115534-G variant was imputed in 230 cases and 182 controls of Amerindian and indigenous ancestry (MAF = 0.027; P = 0.43). Notably, linear regression analyses versus genomic admixture revealed that rs3115534-G was positively correlated with percentage of African ancestry (BETA = 0.064, SE = 0.024, P = 0.01), confirming an African founder effect. At consensus genotyped variants, haplotype size at the *GBA1* risk locus spanning the rs3115534 variant substantially differed across populations when comparing African, African admixed and European PD cases from the *GP2* initiative (European haplotype length = 79.19, European N SNPs = 90; African haplotype length = 19.30, African N SNPs = 29; African admixed haplotype length = 15.15, African admixed N SNPs = 22). Interestingly, the larger sub-African population haplotypes spanning the rs3115534 variant were found in the Esan and the Yoruba in Ibadan (Nigerian) populations according to 1000 Genomes **(Supplementary Figure 9),** suggesting that this haplotype might have originated in these populations, given that founder effects result in decreased genetic diversity and therefore larger haplotype block sizes. Fine-mapping analyses showed the lead SNP had a PP of 71.4% (rs3115534; **Supplementary Table 4).**

### Short- and long-read whole genome sequencing did not identify any coding or structural variant explaining the novel signal at *GBA1*

In an effort to identify a functional coding variant undetectable through genotyping or imputation that could explain the novel GWAS signal, we conducted WGS short-read analyses on a total of 206 individuals (141 cases and 65 controls) of which 39 individuals were *GBA1* rs3115534-GG carriers, 69 were rs3115534-GT and 98 were rs3115534-TT carriers. A 96.6 % correlation was observed between WGS-short read and imputed genotyped data for rs3115534, validating the high quality of our imputed data. No differences in coding variation were observed between carriers and non-carriers of the GWAS signal **(Table 2).** We next applied the Gauchian algorithm, a targeted variant caller for the *GBA1* gene based on WGS BAM files. Gauchian aims to solve the problems caused by the high sequence similarity with the pseudogene paralog *GBAP1 (see methods)*. The Gauchian algorithm did not identify any genetic rearrangement that could explain this signal. Then, Oxford Nanopore Technologies (ONT) WGS long-read sequencing data was generated for a total of five rs3115534-GG PD cases, two rs3115534-GT and six rs3115534-TT controls. Long-read data was compared to short-read WGS for a known structural variant carrier that was previously reported in African American populations in 2000 by Tayebi and colleagues **(Supplementary Figure 10)**^47^. No structural variants explaining this signal were identified. Splice prediction tools (www.phenosystems.com) predicted no significant impact on normal splicing.

**Table 2.**
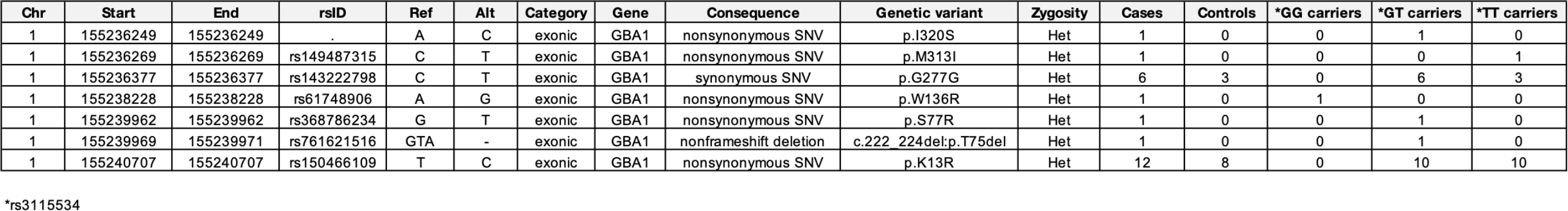
*GBA1* variants identified through Whole Genome Sequencing

### Expression quantitative trait locus analysis provides novel mechanistic insights into risk at the *GBA1* locus

We leveraged existing whole blood expression quantitative trait locus (eQTL) summary statistics from Mak et al., 2021 based on RNA sequencing from 2,733 samples of predominantly African American and Indigenous American ancestries^48^. Of note, we identified a strong eQTL signal at rs3115534, located 8,821 bp from the canonical transcription start site (**Figure 5;** MAF = 0.15; P= 9.99E-25, BETA = 0.238, SE = 0.022). The rs3115534-G risk allele was found to be associated with increased *GBA1* expression levels. We questioned whether this observation could be explained by the existence of multi-mapping reads between *GBA1* and its pseudogene, *GBAP1*, which are often discarded in standard processing and do not contribute to gene-level quantification of expression in many publicly available datasets like GTEx (https://gtexportal.org/). Gustavsson et al., reported that only 42% of all reads mapping to *GBA1* did so uniquely, with the remaining reads mapping primarily to *GBAP1*^49^. This resulted in a significant misestimation of the relative expression of *GBA1* to *GBAP1.* The authors demonstrated the ability of these transcripts to generate stable protein that lacked lysosomal GCase function, which would support our hypothesis. Indeed, transcript diversity is a common and known biological phenomena that could explain the fact that rs3115534-G may increase the expression of a non-functional transcript that in turn would decrease the levels of the transcript responsible for optimal production of the protein isoform with GCase activity. Our data suggests a decreasing trend in GCase activity estimates when comparing rs3115534-GG homozygous risk allele (762.50 ± 273.50 U) versus rs3115534-GT heterozygous carriers (2743.76 ± 1960.83 U); (Welch Two Sample t-test - GG versus GT; t = -4.3138, df = 21.583, p-value = 0.00029) and rs3115534-TT homozygous non-risk allele carriers (1879.94 ± 1010.84 U) versus rs3115534-GG homozygous risk allele carrier; (Welch Two Sample t-test - GG versus TT; t = -4.7564, df = 18.363, p-value = 0.00014). Furthermore, in PD cases alone, the trend in GCase activity between rs3115534-GG homozygous risk allele carriers (762.50 ± 273.50 U), rs3115534-GT heterozygous carriers (3749.47 ± 2620.82 U) and rs3115534-TT homozygous non-risk allele carriers (1976.20 ± 1415.99 U) remained consistent with rs3115534-GG homozygous risk allele displaying the lowest activity; (Welch Two Sample t-test: GG versus GT; t = -3.189, df = 7.3002, p-value = 0.01446; GG versus TT; t = -2.8158, df = 13.003, p-value = 0.01458; GT versus TT; t = 1.7509, df = 9.7545, p-value = 0.1113). All samples were screened for known *GBA1* pathogenic mutations that could bias these estimates. A total of two carriers (one heterozygous for *GBA1* p.I320S and one heterozygous for *GBA1* p.T75del) were removed from our analyses. We assume the limitation that LCLs were only available for one homozygous risk allele. Further research is needed to corroborate this hypothesis and understand the functional consequences of this variant in disease etiology **(Supplementary Figure 11).**

**Figure 5.**
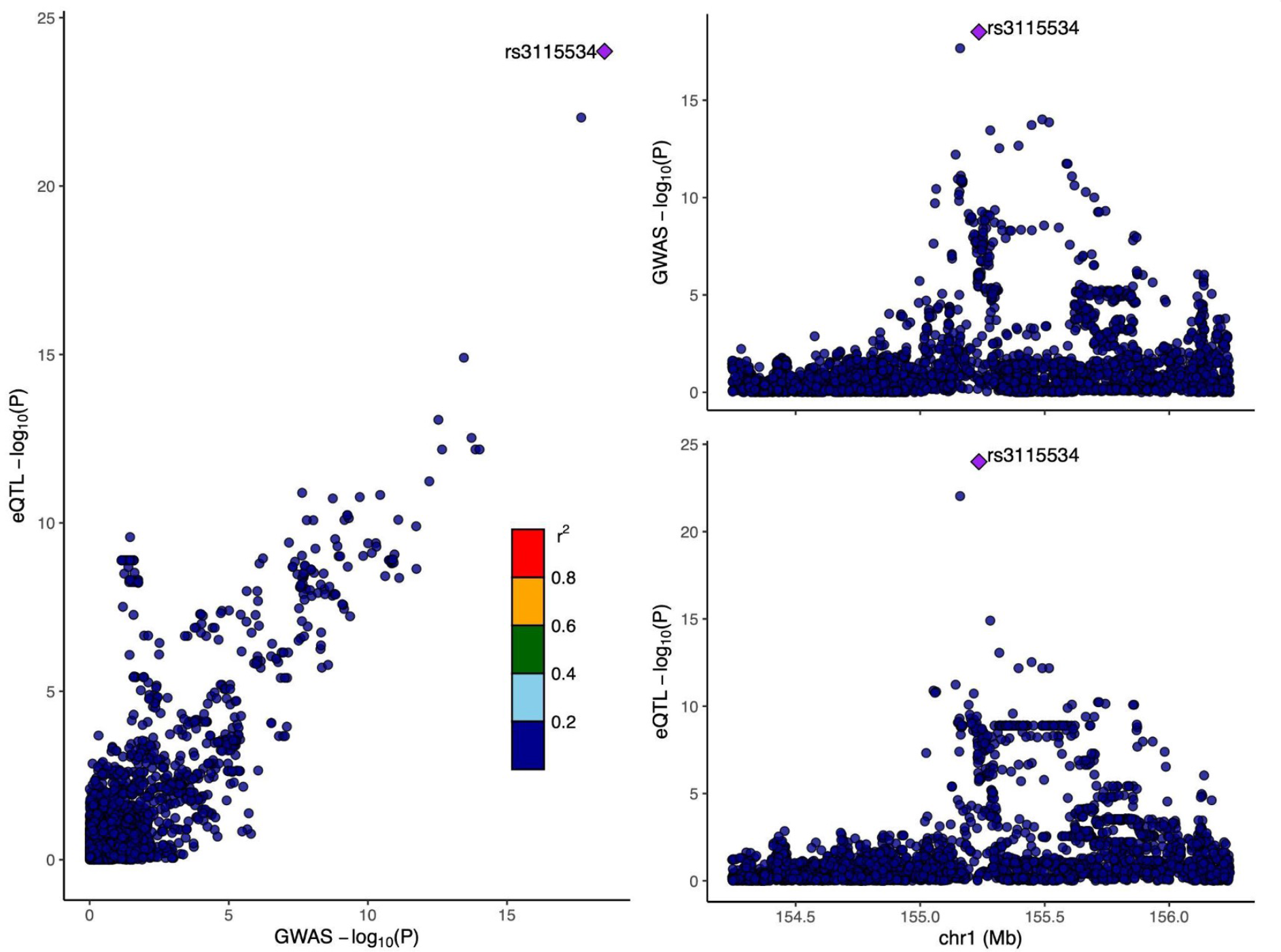
Locus Zoom plot displaying African and African Admixed Parkinson’s disease GWAS Meta-analysis summary statistics versus African expression quantitative trait locus summary statistics from blood (by Mak et al., 2021)

### Characterization of PD known risk loci and polygenic risk profiling suggests some overlapping genetic etiology between European individuals and African and African Admixed populations

The largest PD-GWAS and multi-ancestry GWAS meta-analyses to date identified a total of 104 independent significant PD risk variants ^4,6,8^. Out of the 104 variants, 91 variants passed QC, imputation filters, and were present in the African and African admixed GWAS meta-analysis **(Figure 6**, **Table 2).** Out of the 91 variants, 16 variants were nominally significant (p < 0.05; **Supplementary Table 5**) in the African and African admixed meta-GWAS reported here. We accept the limitation that 23andMe data used in this study was also similar to the data used in the Kim et al., 2022 multi-ancestry GWAS meta-analysis and will have biased our estimates.

**Figure 6.**
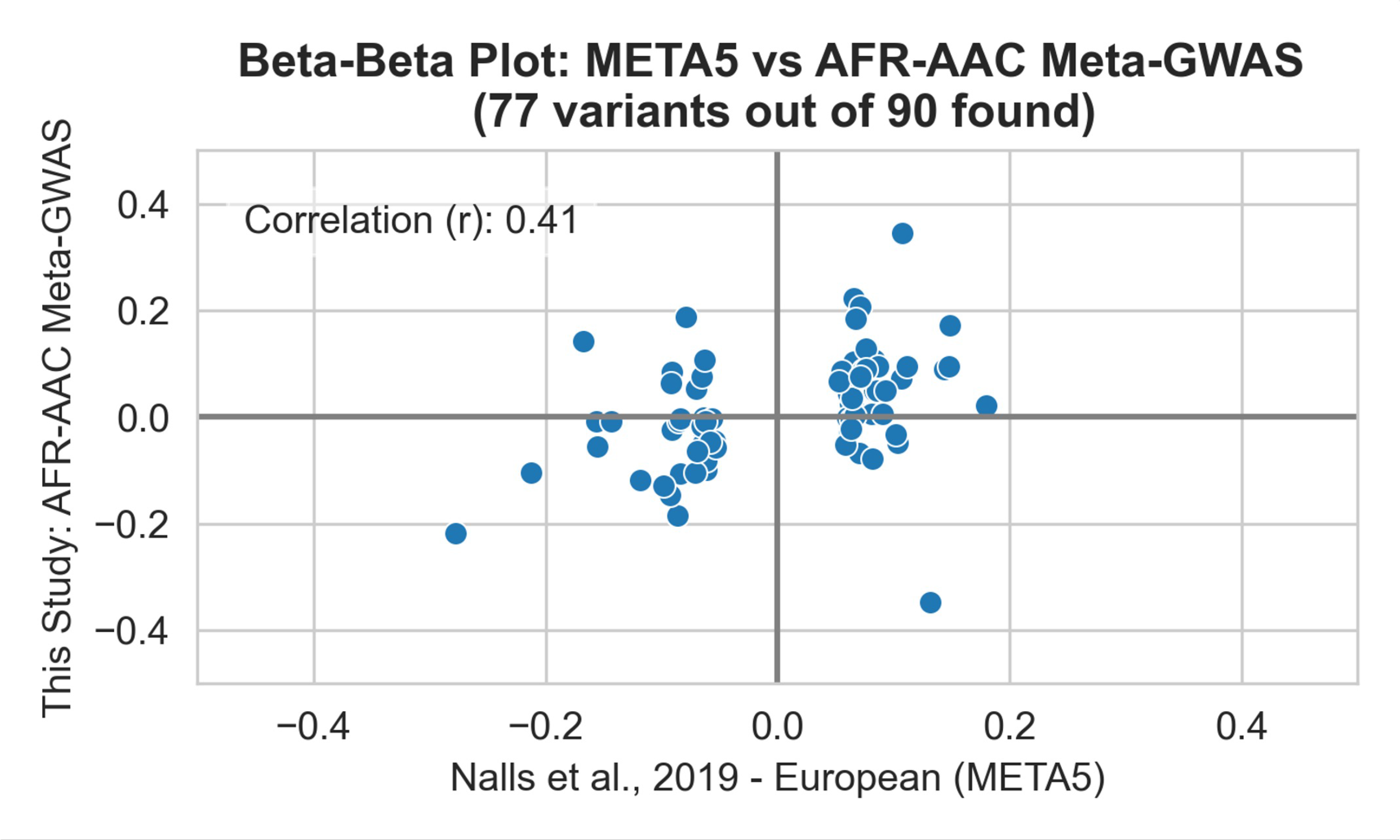
Beta-beta plot comparison of African versus African Admixed estimates for PD known risk loci identified in Europeans

We calculated a PRS using effect estimates from Nalls et al., 2019 summary statistics for the 90 SNPs previously associated with PD risk in European populations. Out of the 90 risk SNPs, a total of 86 passed QC in the African admixed individual level data. European PRS predicted disease status between PD and controls of African admixed ancestry (OR=1.43; 95% CI =1.26-1.61, P=4.37E-05; **Figure 7A**). We then calculated PRS on African individual level data. Out of the 90 risk loci, a total of 79 variants passed QC in the African dataset. African derived PRS predicted disease status between PD and healthy controls of African ancestry (OR=1.27; 95% CI =1.16-1.38, P=1.05E-05; **Figure 7B**). A slightly larger magnitude of effect was observed in the African admixed ancestry PRS model which may be explained by the larger number of SNPs that passed QC or larger percentage of admixed European ancestry within this cohort. The variant with the largest effect size in both models was chr4:89704960:G:A_A (rs356182), located at the *SNCA* locus. After adjusting for rs356182, PRS differences between PD and controls were still significant (PRS_African admixed_ P =2.21E-05 ; PRS_African_ P =0.014). As expected, the observed effect is consistently lower as compared to Europeans (PRS OR_training dataset_ =3.74, 95% CI =3.35 –4.18)^4^ suggesting that additional novel genetic loci might contribute to the heritability of PD on the African and African admixed populations.

**Figure 7.**
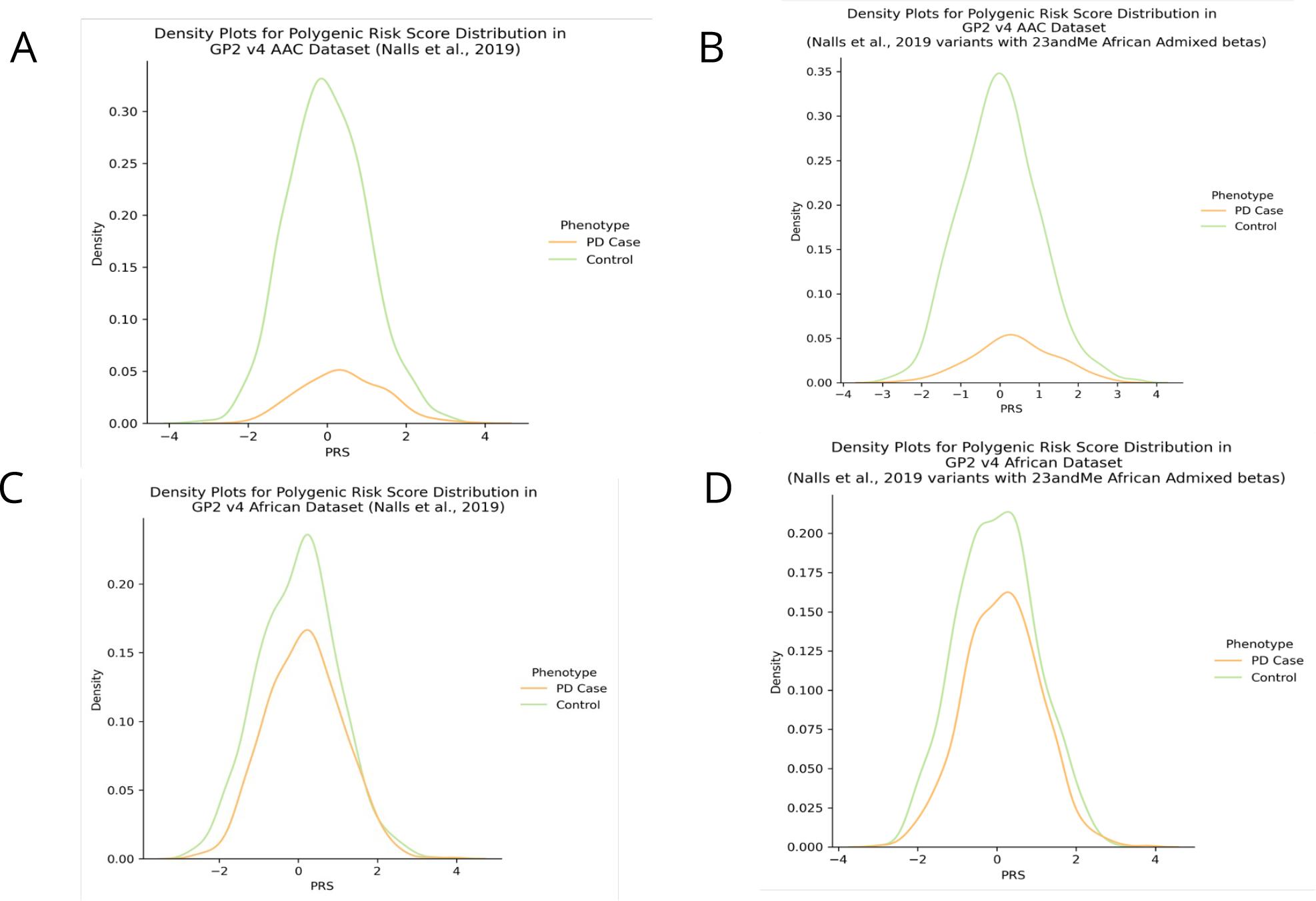
Density plots showing polygenic risk score distributions in the African and African Admixed individuals using the 90 Parkinson’s disease risk loci

Furthermore, we calculated a PRS using effect estimates from the AAC 23andMe summary statistics *(see **Methods**)* for the same 90 SNPs previously associated with PD risk. Out of the 90 risk SNPs, a total of 77 passed QC in the African and African admixed individual level datasets from *GP2*. PRS predicted disease status between PD and healthy controls of African admixed ancestry (OR=1.26; 95% CI =1.15-1.37; P=1.89E-05; **Figure 7C**). We then calculated PRS on African individual level data. PRS predicted disease status between PD and healthy controls of African ancestry (OR=1.42; 95% CI =1.25-1.60; P=6.65E-05; **Figure 7D**). When using AAC 23andMe summary statistics, the PRS effect size was slightly higher when applied to African individual level data than when using Nalls et al., 2019 European summary statistics as the reference dataset.

### Enrichment of homozygosity patterns in African and African Admixed cases suggests yet to be discovered recessive variants linked to Parkinson’s disease etiology

Although no enrichment of specific ROH was identified spanning the *GBA1* locus, we identified longer runs of homozygosity in cases compared to controls at a genome-wide level. The number of segments was associated with PD risk at an OR of 1.08 per 1Mb increase in average number of ROH (OR = 1.08; 95% CI = 1.066- 1.11; P = 4.63E-16). The total size of these homozygosity regions were also found to be significantly different between cases and controls, although with a small effect (BETA = 1.69E-05 per kb increase; SE = 3.531E-06; P = 1.53E-06). This suggests that yet to be uncovered recessive variants might, to some extent, contribute to PD etiology in these populations.

## Discussion

There have been only a few published studies exploring PD genetics in the African and African admixed populations, conducted with fewer than thirty samples in all instances^50–54^. In the present study, we have gathered the largest collection of PD patients and controls from African and African admixed ancestry populations to comprehensively assess the genetic architecture of PD on a genome-wide scale. Here, we identified a novel African-specific GWAS signal on the *GBA1* locus, significantly associated with PD risk and AAO, to be the most important risk factor for PD in this African and African admixed populations. In contrast, initial well powered GWAS in European populations nominated the *SNCA* and *MAPT* loci as the most significant contributors to PD genetic risk in Europeans. Remarkably, almost a four times larger sample size in cases was required to nominate *GBA1* as one of the major PD risk factors in the European ancestry population through GWAS,^55^ showing the power and benefit of using diverse ancestry data.

We suggest a novel disease mechanism via expression changes consistent with a trend towards decreased GCase activity levels. The *GBA1* c.1225-34C>A (rs3115534) GWAS hit alters a non-conserved intronic nucleotide (GERP++ score = -2.04). Despite the large effect size driven by this signal, our study did not identify an association with any previously reported or new *GBA1* coding or structural aberration that could explain this signal^39, 47, 56, 57^. Splice prediction tools predicted no significant impact on normal splicing, while rs3115534 has been reported to be an expression quantitative trait locus (eQTL) in several tissues^48, 58^. Additionally, a large-scale pQTL study in African Americans with chronic kidney disease suggests that at the protein level the risk allele for PD in our GWAS (G) is associated with a reduction in the level of GCase protein in blood, as defined by the SOMAscan assay. This finding supports the concept that the risk allele leads to a partial loss of both GCase protein and GCase enzyme activity^59^.

Strikingly, by leveraging existing eQTL data predominantly of African American ancestry, we found the rs3115534-G risk allele to be associated with increased *GBA1* expression levels in whole blood, but paradoxically linked with a trend towards decreased GCase activity, which may be due to challenges with RNA-seq in this locus. This interesting finding, possibly explained by transcript diversity leading to the expression of a protein with diminished lysosomal GCase activity, warrants further study. Previously, *GBA1* variants associated with PD risk have all been coding mutations, but here we identify a novel functional mechanism involved in disease etiology. Our findings are limited by the absence of brain QTL data in non-European populations, underscoring the importance of increasing representation from ancestrally diverse populations to enable new discoveries and ensure their equitable translation. Future large scale single cell expression studies should investigate in which brain cell types these expression differences are most prominent. This novel mechanism opens new avenues towards efficient RNA-based therapeutic strategies, such as antisense oligonucleotides or short interfering RNAs aimed at reducing lifetime risk.

Interestingly, given the high population frequency of the identified signal and the phenotypic characteristics of the homozygous Africans and African admixed carriers, our study does not support the notion that this variant causes Gaucher disease. Furthermore, the rs3115534 variant has been found to be extremely rare in non-African/African admixed populations. These findings suggest an African founder effect, and reinforce that the genetic architecture of this locus and its influence in risk and onset is different across populations. Interestingly, rs3115534 was also found to be associated with PD AAO in our study. The largest GWAS meta-analysis investigating the role of genetic determinants on PD onset in European populations^60^ nominated p.N409S as an AAO disease modifier. This variant, which is one of the most common *GBA1* risk factors in European and Ashkenazi Jewish populations, is 100 times less frequent in individuals of African and African admixed ancestry. In support of this notion, we did not find any of the common *GBA1* pathogenic variants through WGS in this study.

PRSs predicted disease status between predominantly PD cases and healthy controls of African and African admixed ancestry, with a slightly better prediction when using African admixed summary statistics on African individual level data. Interestingly, *SNCA*-rs356182, the lead risk variant at this locus in European and Amerindian and indigenous ancestry studies, was found to be the major genetic player in both models ^4, 7^. This finding is in concordance with a partial overlapping directionality of effect observed between PD known risk alleles predisposing to disease in European and African populations. While this is a start, additional data and understanding of the disease in other African sub-populations is needed to improve prediction models per ancestry. Our current study used effect estimates from 288 African admixed PD cases and 193,985 controls provided by 23andMe. As population-specific studies become better powered for each of these sub-populations, we believe effect estimates and, in turn, predictions will improve. Intriguingly, we revealed an overall excess of homozygosity in PD cases versus controls, pointing out the possible existence of disease-causing recessive variants that might be uncovered by future sequencing analysis in these populations.

Although we have made progress in assessing genetic risk factors for PD in an African-specific manner, there are a number of limitations to our study. Unraveling additional susceptibility genetic risk and phenotypic relationships would have been possible if a larger cohort had been analyzed. Additionally, due to lack of well-powered and African-specific RNA sequencing datasets and the added complexity of multi-mapping reads between *GBA* and *GBAP1*, we assume the limitation that this potential novel functional mechanism merits further study. While this study marks major progress in assessing genetic risk factors for PD, there remains a great deal to be done. Future studies should explore the effect of this variant on cognitive impairment in PD.

Here, we produce crucial insights into targeted construction of African ancestral haplotypes and potential novel pathogenic mechanisms underlying PD etiology. The utility of genetically characterizing populations of African and African admixed ancestry is unquestionable^61^. This study demonstrates the importance of haplotype substructure discoveries for future fine-mapping efforts, showing how leveraging unique populations can benefit our understanding of complex diseases.

Overall, addressing the genetic complexity underlying these underrepresented populations, our study represents a valuable resource for identifying and tracking *GBA1* carriers that may prove relevant for enrollment in target-specific PD clinical trials. *GBA1* genetic testing in the African and African admixed populations can help to design an optimized trial with the highest likelihood of providing meaningful results and actionable answers. We envisage that these data generated under the Global Parkinson’s Genetics Program initiative will be key to shed light on the molecular mechanisms involved in the disease process and might pave the way for future clinical trials and therapeutic interventions.

This would be helpful to further improve our granularity in association testing and ability to fine-map through integration of omics data while also evaluating population specific associations.

## Data Sharing

All GP2 data is hosted in collaboration with the Accelerating Medicines Partnership in Parkinson’s disease, and is available via application on the website (https://amp-pd.org/register-for-amp-pd). The GWAS summary statistics from this study, excluding 23andMe, are available as of GP2’s release 5.

23andMe summary statistics are available via application on the website (https://research.23andme.com/dataset-access/). Genotyping imputation, quality control, ancestry prediction, and processing was performed using GenoTools v1.0, publicly available on GitHub (https://github.com/GP2code/GenoTools). All scripts for analyses are publicly available on GitHub [https://github.com/GP2code/GP2-AFR-AAC-metaGWAS; 10.5281/zenodo.7888141].

## Ethics Statement

All cohorts recruited to the GP2 initiative undergo a thorough review of the consent forms in the Operations and Compliance working group, ensuring that each contributing study abided by the ethics guidelines set out by their institutional review boards, and all participants gave informed consent for inclusion in both their initial cohorts and subsequent studies within local law constraints. All GP2 data is hosted in collaboration with the Accelerating Medicines Partnership in Parkinson’s disease, and is available via application on the website (https://amp-pd.org/register-for-amp-pd).

Summary statistics for individuals with or without PD were provided through a collaborative agreement with 23andMe, Inc. Participants provided informed consent and volunteered to participate in the research online, under a protocol approved by the external AAHRPP-accredited IRB, Ethical & Independent (E&I) Review Services. As of 2022, E&I Review Services is part of Salus IRB (https://www.versiticlinicaltrials.org/salusirb). 23andMe summary statistics are available via application on the website (https://research.23andme.com/dataset-access/).

## Declaration of Interests

This research was supported in part by the Intramural Research Program of the NIH, National Institute on Aging (NIA), National Institutes of Health, Department of Health and Human Services; project number ZO1 AG000535 and ZIA AG000949, as well as the National Institute of Neurological Disorders and Stroke (NINDS) and the National Human Genome Research Institute (NHGRI).

This work was supported in part by the Global Parkinson’s Genetics Program (GP2). GP2 is funded by the Aligning Science Against Parkinson’s (ASAP) initiative and implemented by The Michael J. Fox Foundation for Parkinson’s Research (https://gp2.org). Additional funding was provided by The Michael J. Fox Foundation for Parkinson’s Research through grant MJFF-009421/17483. For a complete list of GP2 members see https://gp2.org D.V., H.I., H.L.L., K.L. and M.A.N.’s participation in this project was part of a competitive contract awarded to Data Tecnica International LLC by the National Institutes of Health to support open science research.

K.H. and members of the 23andMe Research Team are employed by and hold stock or stock options in 23andMe, Inc. M.A.N. also currently serves on the scientific advisory board for Character Biosciences Inc and Neuron 23 Inc.

DGS is a member of the faculty of the University of Alabama at Birmingham and is supported by endowment and University funds, is an investigator in studies funded by Abbvie, Inc., the American Parkinson Disease Association, the Michael J. Fox Foundation for Parkinson Research, The National Parkinson Foundation, Alabama Department of Commerce, Alabama Innovation Fund, Genentech, the Department of Defense, and NIH grants P50NS108675 and R25NS079188 and has a clinical practice and is compensated for these activities through the University of Alabama Health Services Foundation. He serves as Deputy Editor for the journal Movement Disorders and is compensated for this role by the International Parkinson and Movement Disorders Society. In addition, since January 1, 2022 he has served as a consultant for or received honoraria from Abbvie Inc., Curium Pharma, Appello, Theravance, Sanofi-Aventis, Alnylam Pharmaceutics, Coave Therapeutics, BlueRock Therapeutics and F. Hoffman-La Roche.

We thank the research participants and employees of 23andMe. The following members of the 23andMe Research Team contributed to this study: Stella Aslibekyan, Adam Auton, Elizabeth Babalola, Robert K. Bell, Jessica Bielenberg, Katarzyna Bryc, Emily Bullis, Paul Cannon, Daniella Coker, Gabriel Cuellar Partida, Devika Dhamija, Sayantan Das, Sarah L. Elson, Nicholas Eriksson, Teresa Filshtein, Alison Fitch, Kipper Fletez-Brant, Pierre Fontanillas, Will Freyman, Julie M. Granka, Alejandro Hernandez, Barry Hicks, David A. Hinds, Ethan M. Jewett, Yunxuan Jiang, Katelyn Kukar, Alan Kwong, Keng-Han Lin, Bianca A. Llamas, Maya Lowe, Jey C. McCreight, Matthew H. McIntyre, Steven J. Micheletti, Meghan E. Moreno, Priyanka Nandakumar, Dominique T. Nguyen, Elizabeth S. Noblin, Jared O’Connell, Aaron A. Petrakovitz, G. David Poznik, Alexandra Reynoso, Madeleine Schloetter, Morgan Schumacher, Anjali J. Shastri, Janie F. Shelton, Jingchunzi Shi, Suyash Shringarpure, Qiaojuan Jane Su, Susana A. Tat, Christophe Toukam Tchakouté, Vinh Tran, Joyce Y. Tung, Xin Wang, Wei Wang, Catherine H. Weldon, Peter Wilton, Corinna D. Wong We thank Cynthia J. Casaceli, Debbie Baker and Christi Alessi-Fox from the University of Rochester Clinical Trials Coordination Center for its contribution to the coordination of the BLAAC PD Study. We thank Lisa Shulman for her contribution to the design of the protocol for BLAAC PD clinical assessments.

We thank the Biowulf team, as this study used the high-performance computational capabilities of the Biowulf Linux cluster at the National Institutes of Health (http://hpc.nih.gov).

Figure 1 was generated on www.biorender.com.

## Data Availability

All GP2 data is hosted in collaboration with the Accelerating Medicines Partnership in Parkinsons disease, and is available via application on the website (https://amp-pd.org/register-for-amp-pd). The GWAS summary statistics from this study, excluding 23andMe, are available as of GP2 release 5. 23andMe summary statistics are available via application on the website (https://research.23andme.com/dataset-access/). Genotyping imputation, quality control, ancestry prediction, and processing were performed using GenoTools v1.0, publicly available on GitHub (https://github.com/GP2code/GenoTools). All scripts for analyses are publicly available on GitHub [https://github.com/GP2code/GP2-AFR-AAC-metaGWAS; 10.5281/zenodo.7888141].

https://research.23andme.com/dataset-access/

https://github.com/GP2code/GenoTools

https://github.com/GP2code/GP2-AFR-AAC-metaGWAS

## Supplementary Tables

**Supplementary Table 1.** Genome-wide significant SNPs identified in the African only GWAS meta-analysis

**Supplementary Table 2.** Genome-wide significant SNPs identified in the African and African Admixed GWAS meta-analysis

**Supplementary Table 3.** Allele frequencies for *GBA1* - rs3115534 in African and African admixed subpopulations

**Supplementary Table 4.** Meta-GWAS fine-mapping analyses

**Supplementary Table 5.** Genome-wide replication assessment of known PD risk loci.

**Supplementary Table 6.** Long-Read Whole Genome Sequencing statistics

**Supplementary Table 7.** The 23andMe Reference Panel Information

