## Supplemental Methods and Figures for "Genome-wide Association Identifies Novel Etiological Insights Associated with Parkinson’s Disease in African and African Admixed Populations"

*joint first
[#]joint last
[CA]Corresponding

*Tables (see Supplementary_Tables.xlsx)*

**Supplementary Table 1.** Genome-wide significant SNPs identified in the African only GWAS meta-analysis ........ ST1

**Supplementary Table 2.** Genome-wide significant SNPs identified in the African and African Admixed GWAS meta-analysis ........ ST2

**Supplementary Table 3.** Allele frequencies for *GBA1 - rs3115534* in African and African admixed subpopulations ........ ST3

**Supplementary Table 4.** Meta-GWAS fine-mapping analyses ........ ST4

**Supplementary Table 5.** Genome-wide replication assessment of known PD risk loci. ........ ST5

**Supplementary Table 6.** Long-Read Whole Genome Sequencing statistics ........ ST6

**Supplementary Table 7.** The 23andMe Reference Panel Information ........ ST7

*Methods*

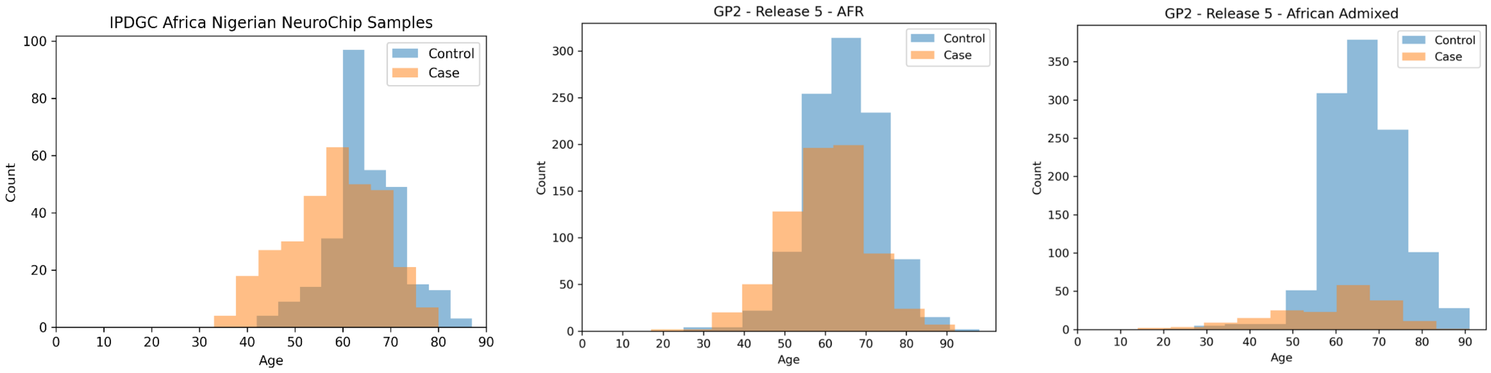

**Supplementary Figure 1** | Age distributions of cohorts under study

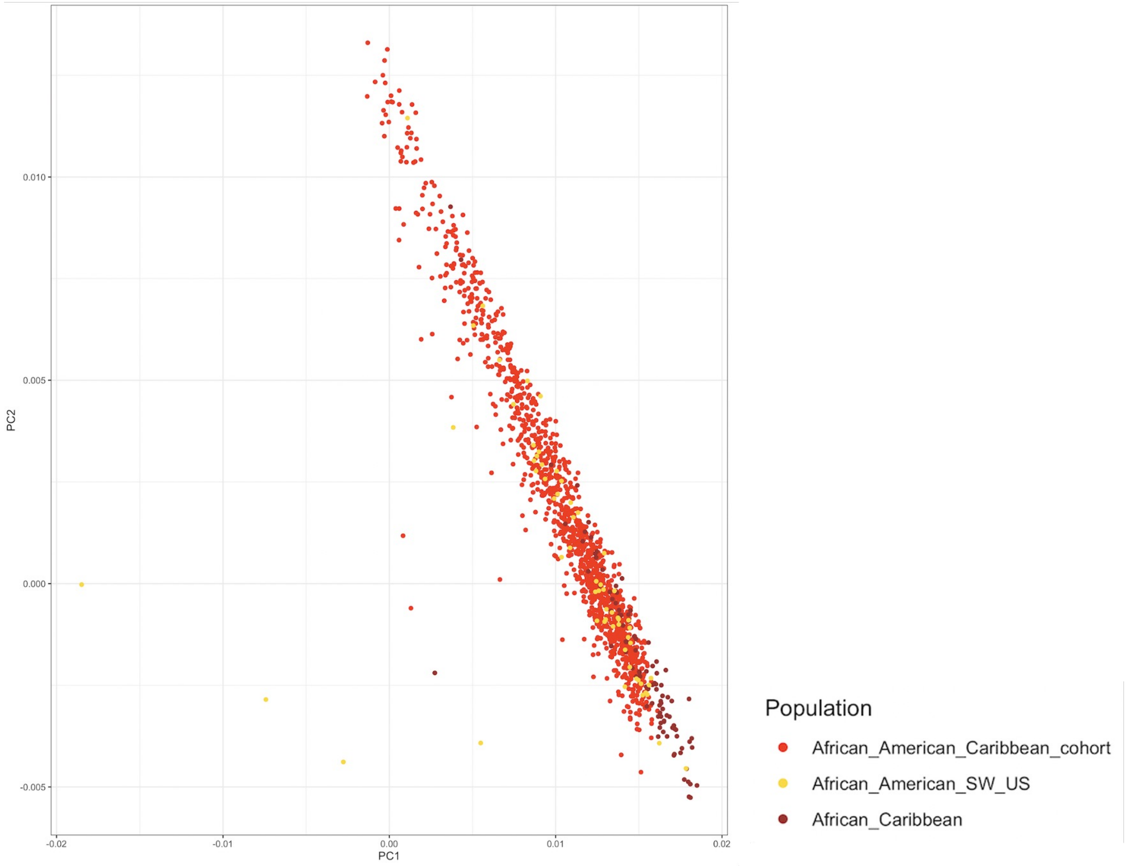

**Supplementary Figure 2** | African Admixed cohort with 1000 Genome populations

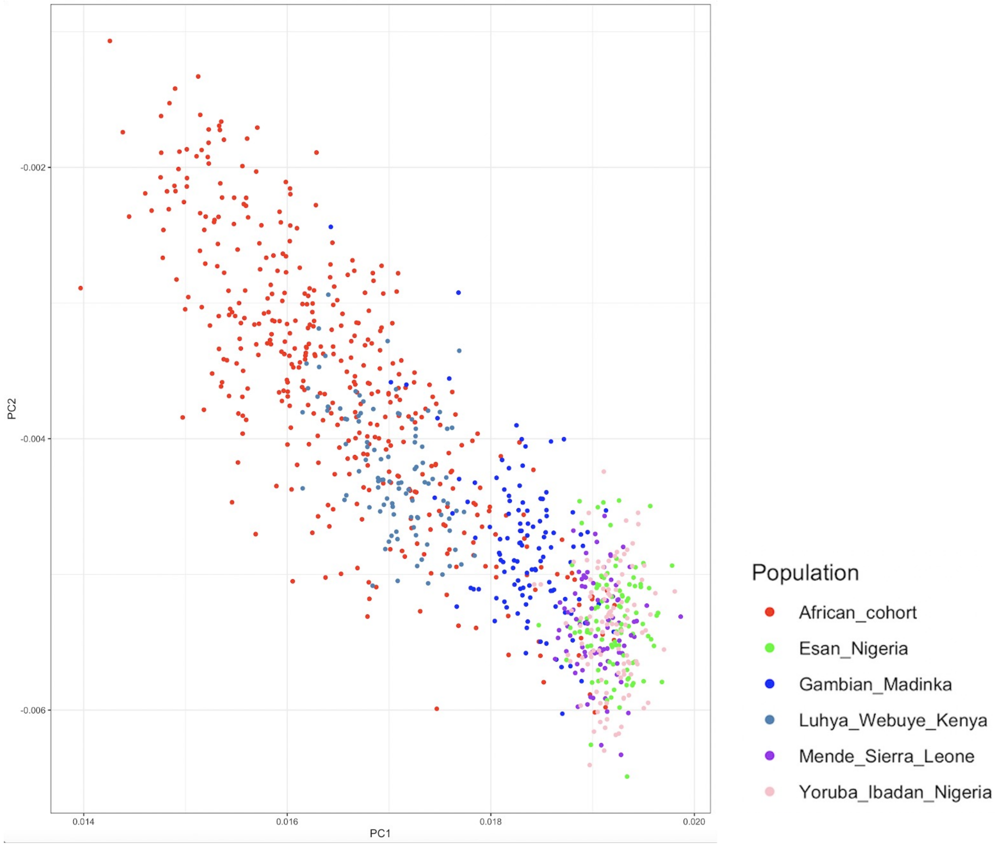

**Supplementary Figure 3** | African cohort with 1000 Genome populations

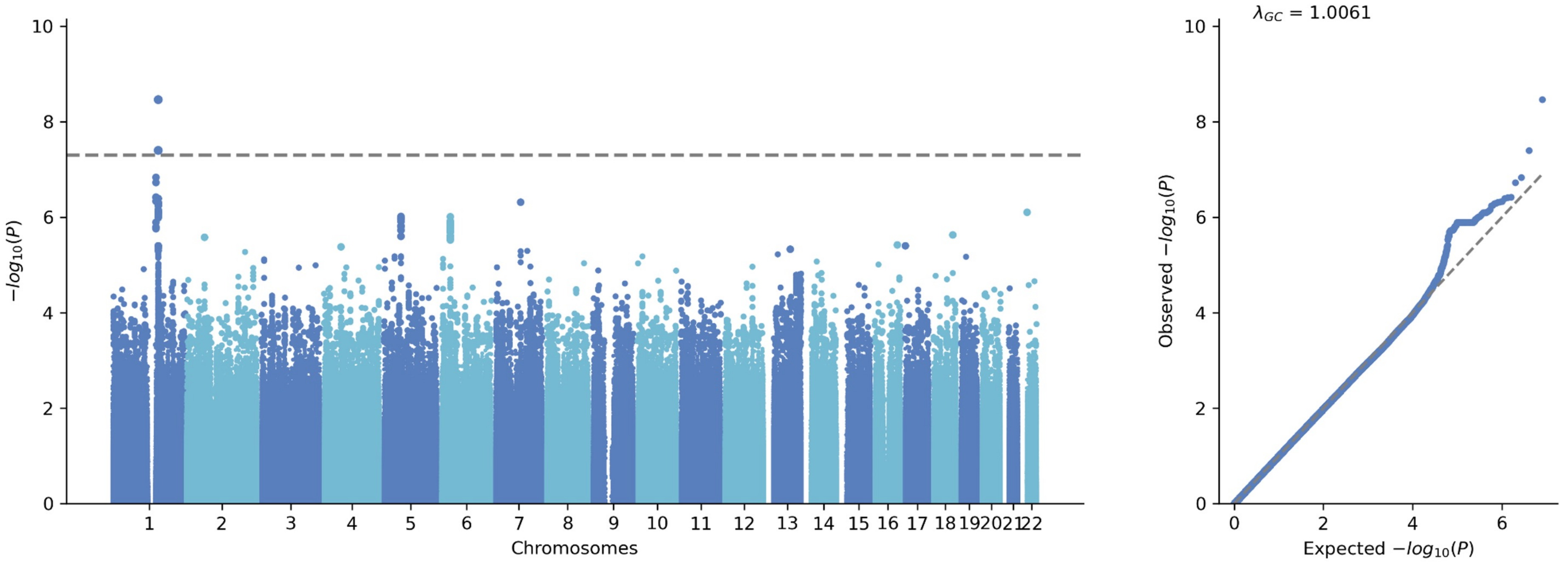

**Supplementary Figure 4** | African Parkinson's disease risk GWAS

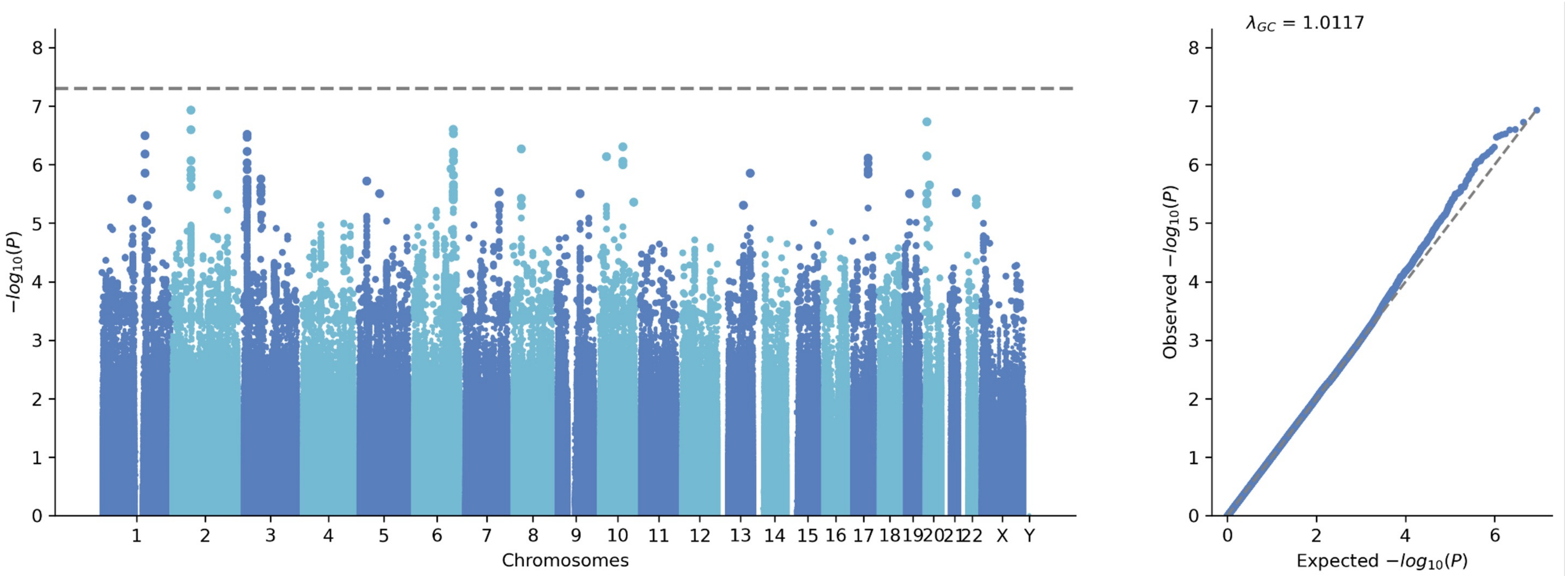

**Supplementary Figure 5** | African Admixed Parkinson's disease risk GWAS

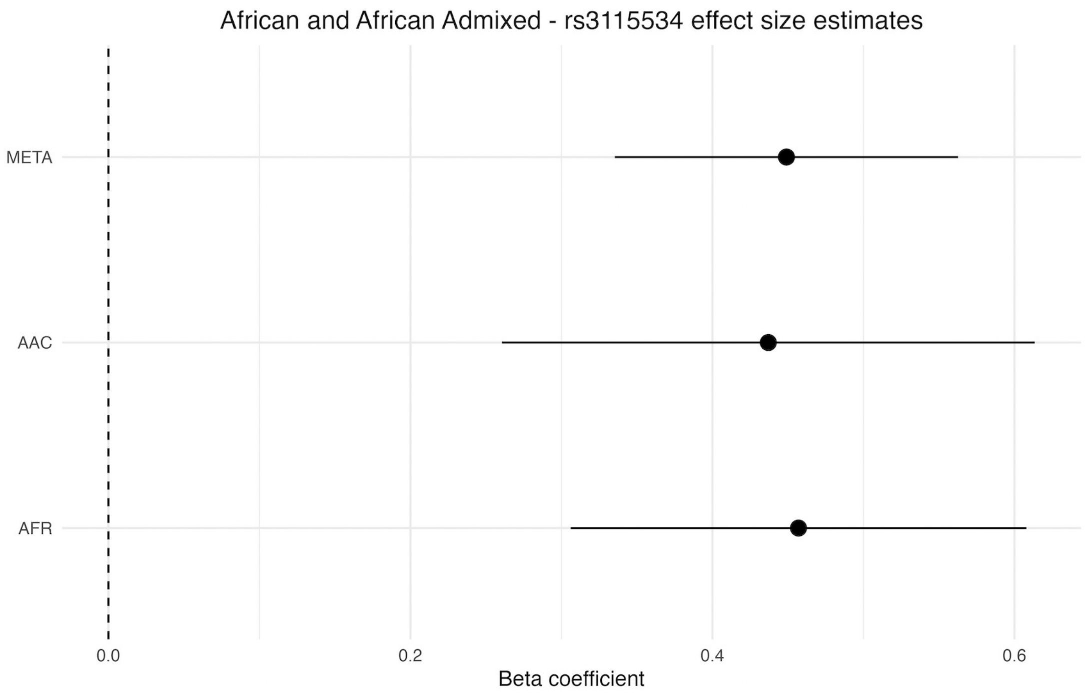

| COHORT | BETA | SE | L95 | U95 | P |
|---|---|---|---|---|---|
| AAC | 0.437 | 0.09 | 0.2606 | 0.6134 | 1.40E-06 |
| AFR | 0.457 | 0.077 | 0.30608 | 0.60792 | 3.44E-09 |
| META | 0.449 | 0.058 | 0.33532 | 0.56268 | 2.40E-14 |

**Supplementary Figure 6**| Forest plot displaying effect size estimates for *GBA* - rs3115534 in the African, African admixed and overall meta-analysis datasets.

The circle denotes the beta coefficient for *GBA* - rs3115534, with the horizontal lines indicating the 95% confidence intervals.

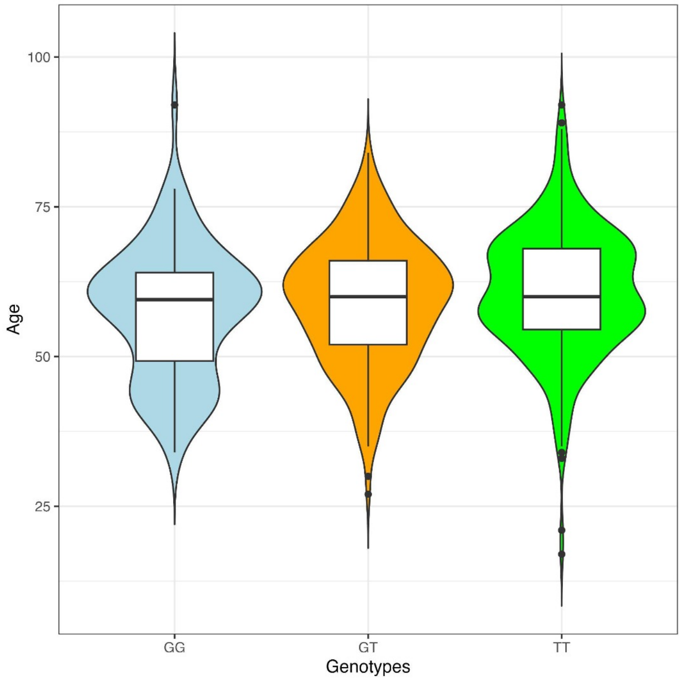

**Supplementary Figure 7** | *GBA1* - rs3115534 Genotypes versus age at Parkinson's disease onset

rs3115534-GG versus age at onset; BETA =-1.96, SE = -0.64, P=0.002; rs3115534-GT versus age at onset; BETA =-2.28, SE =0.85 , P=0.007.

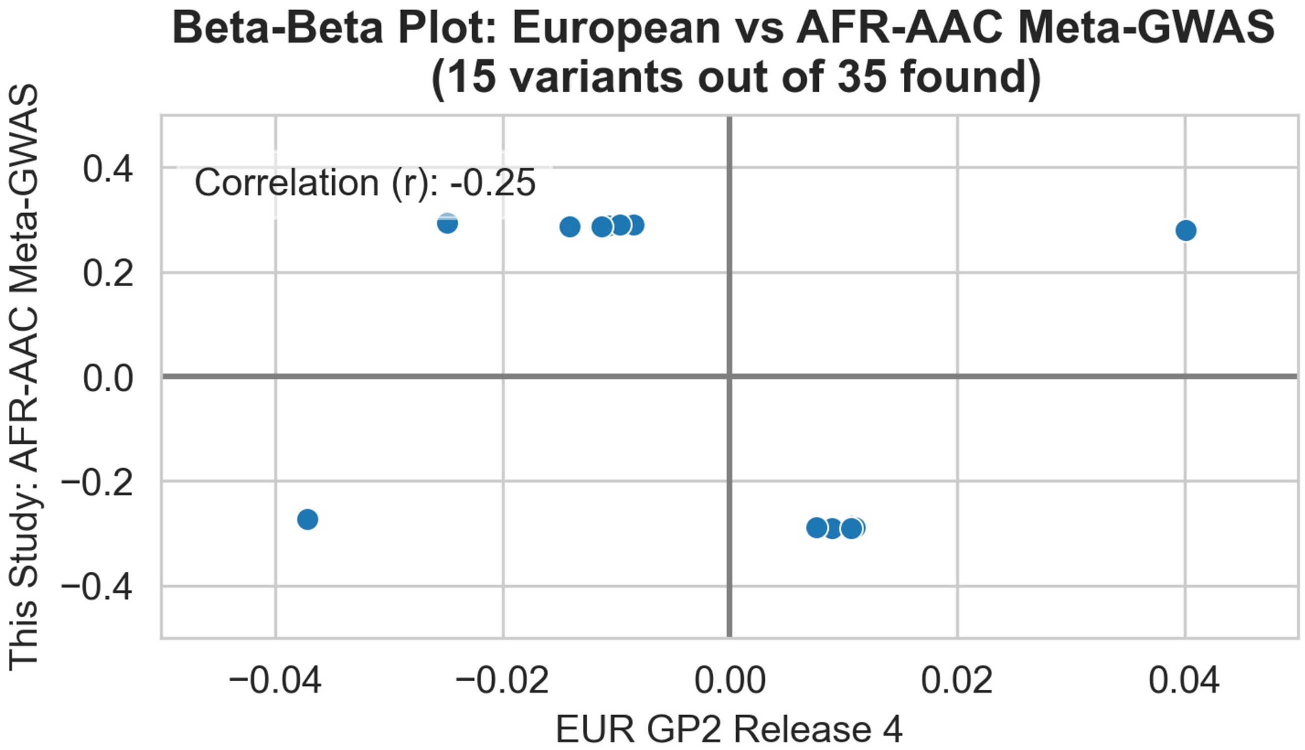

**Supplementary Figure 8** | Beta-beta plot comparison of Europeans for PD *GBA1* risk locus identified in African and African Admixed

**Esan in Nigeria**

| RS Number | Position (GRCh37) | Allele Frequencies | Haplotypes | |
|---|---|---|---|---|
| rs3115534 | chr1:155205669 | T=0.697, G=0.303 | T | G |
| | | Haplotype Count | 138 | 60 |
| | | Haplotype Frequency | 0.697 | 0.303 |

**Yoruba in Ibadan**

| RS Number | Position (GRCh37) | Allele Frequencies | Haplotypes | |
|---|---|---|---|---|
| rs3115534 | chr1:155205669 | T=0.718, G=0.282 | T | G |
| | | Haplotype Count | 155 | 61 |
| | | Haplotype Frequency | 0.7176 | 0.2824 |

**Luhya in Webuye, Kenya**

| RS Number | Position (GRCh37) | Allele Frequencies | Haplotypes | |
|---|---|---|---|---|
| rs3115534 | chr1:155205669 | T=0.823, G=0.177 | T | G |
| | | Haplotype Count | 163 | 35 |
| | | Haplotype Frequency | 0.8232 | 0.1768 |

**Gambian in Western Division**

| RS Number | Position (GRCh37) | Allele Frequencies | Haplotypes | |
|---|---|---|---|---|
| rs3115534 | chr1:155205669 | T=0.827, G=0.173 | T | G |
| | | Haplotype Count | 187 | 39 |
| | | Haplotype Frequency | 0.8274 | 0.1726 |

**Mende in Sierra Leone**

| RS Number | Position (GRCh37) | Allele Frequencies | Haplotypes | |
|---|---|---|---|---|
| rs3115534 | chr1:155205669 | T=0.829, G=0.171 | T | G |
| | | Haplotype Count | 141 | 29 |
| | | Haplotype Frequency | 0.8294 | 0.1706 |

**African Ancestry in Southwest USA**

| RS Number | Position (GRCh37) | Allele Frequencies | Haplotypes | |
|---|---|---|---|---|
| rs3115534 | chr1:155205669 | T=0.885, G=0.115 | T | G |
| | | Haplotype Count | 108 | 14 |
| | | Haplotype Frequency | 0.8852 | 0.1148 |

**African – Caribbean Black**

| RS Number | Position (GRCh37) | Allele Frequencies | Haplotypes | |
|---|---|---|---|---|
| rs3115534 | chr1:155205669 | T=0.802, G=0.198 | T | G |
| | | Haplotype Count | 154 | 38 |
| | | Haplotype Frequency | 0.8021 | 0.1979 |

**Supplementary Figure 9** | Haplotype frequencies spanning *GBA1* - rs3115534 per African and African admixed subpopulation

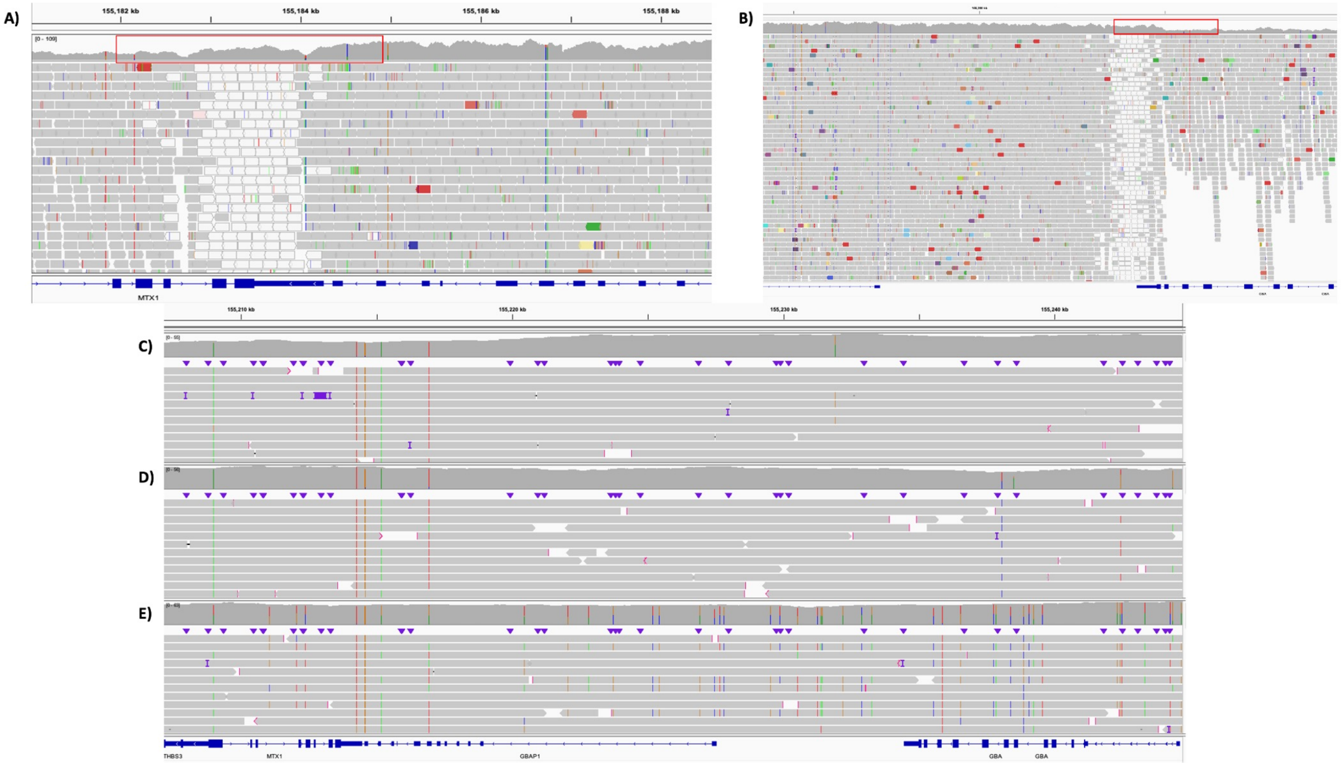

#### Supplementary Figure 10 | *GBAP1* Duplication and MTX1, MTX1P1 Fusion vs Long Read Sequencing

In 2000, Tayebi and colleagues identified a novel recombination occurring more frequently in African American patients and resulting in a duplication of glucosylceramidase beta pseudogene 1 (*GBAP1*) and fusion of metaxin (*MTX1*) and its pseudogene (*MTX1P1*) (PMID: 11129343, PMID: 11241475). Dr. Ellen Sidransky and Dr. Nahid Tayebi generously provided whole genome sequencing data (**A**,**B**) for a sample containing this recombination which was visualized on Broad Institute Integrative Genomics Viewer (2.15.4). We generated long read sequencing data for five rs3115534-GG carriers (5 PD cases), two rs3115534-GT carriers (2 PD cases), and 6 rs3115534-TT carriers (2 PD case; 4 PD controls). A handful of these samples are shown above for reference; **C)** rs3115534-GG (PD case), **D)** rs3115534-GG (PD case), **E)** rs3115534-TT (PD case). The long read sequencing data we generated was compared to the recombination found by Tayebi and colleagues. Coverage changes in A and B are highlighted in red and indicate the presence of a recombination. Our long read sequencing data failed to reveal any structural variants in this region.

#### GCase Activity (U) by Genotype Average (All)

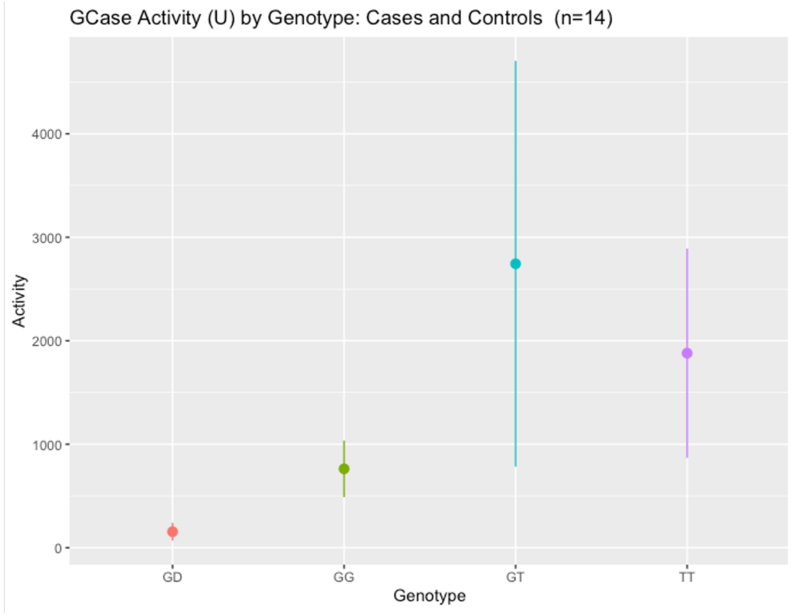

#### GCase Activity (U) per Sample by Genotype (All)

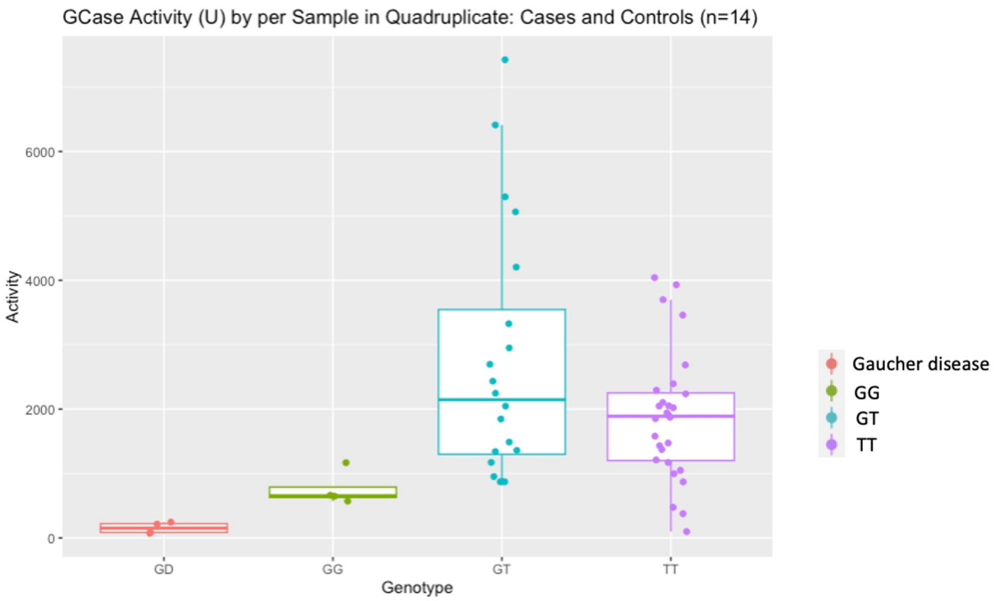

**Supplementary Figure 11** | GCase activity analyses performed on *GBA1* - rs3115534-GG, rs3115534-GT, and rs3115534-TT carriers

A fluorometric 4-MU assay was used to measure GCase activity in 14 lymphoblastoid cell lines, including a type I Gaucher disease (GD) patient as a positive control. **A)** Samples were aggregated by rs3115534 genotype and average activity. Values are represented by mean and standard deviation. **B)** All 14 samples were run in quadruplicate. Samples were screened for known *GBA1* pathogenic mutations that could bias these estimates. A total of two carriers (one heterozygous for *GBA1* p.I320S and one heterozygous for *GBA1* p.T75del) were removed from further analyses.

#### GCase Activity (U) by Genotype Average (PD Only)

#### GCase Activity (U) by Genotype per Sample (PD Only)

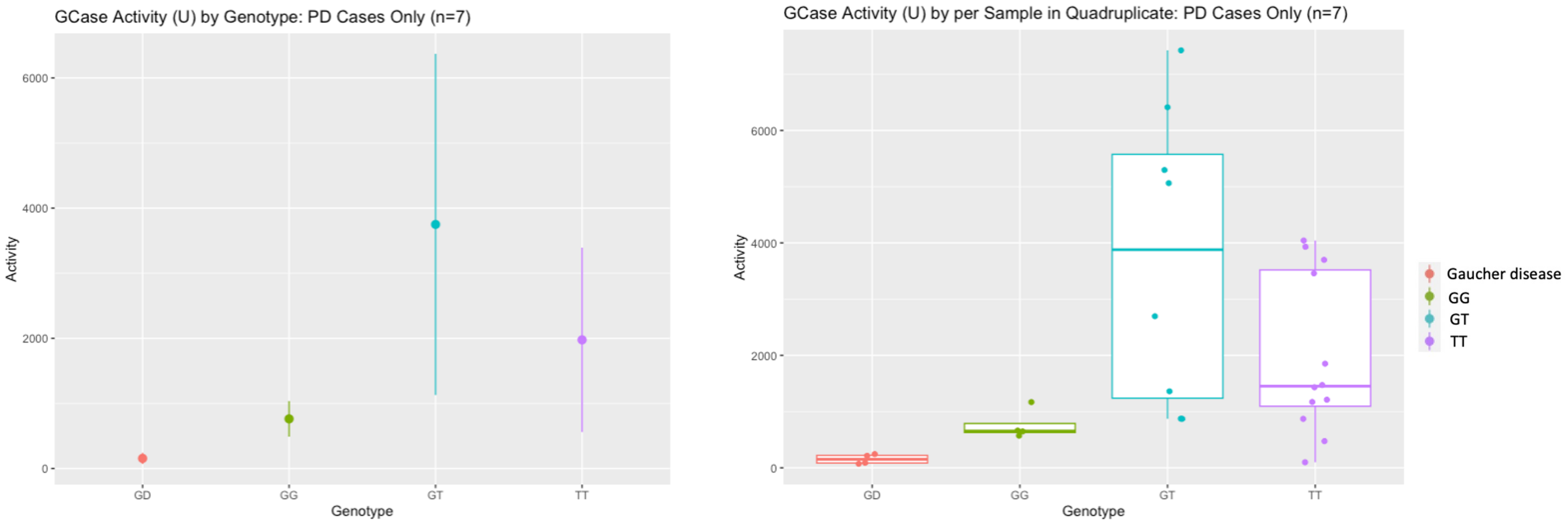

**Supplementary Figure 12** | GCase activity analyses performed on *GBA1* - rs3115534-GG, rs3115534-GT, and rs3115534-TT carriers

Samples with Parkinson’s disease were pulled from the 14 samples shown in Supplementary Figure 11, including a type I Gaucher disease (GD) patient as a positive control. **A)** A total of 9 samples with Parkinson’s disease were aggregated by rs3115534 genotype and average activity. Values are represented by mean and standard deviation. **B)** All 9 samples with Parkinson’s were run in quadruplicate (Welch Two Sample t-test: GG versus GT; t = -3.189, df = 7.3002, p-value = 0.0.01446; GG versus TT; t = -2.8158, df = 13.003, p-value = 0.01458; GT versus TT; t = 1.7509, df = 9.7545, p-value = 0.1113). A total of two carriers (one heterozygous for GBA1 p.I320S and one heterozygous for GBA1 p.T75del) were removed from further analyses.

### Methods

#### GP2 ancestry Prediction

Raw genotype data was passed through an ancestry prediction and pruning machine learning method as a part of the GenoTools pipeline (https://github.com/GP2code/GenoTools), elaborated on elsewhere23. In brief, ancestry was defined using reference panels from the 1000 Genomes Project24, Human Genome Diversity Project25, and an Ashkenazi Jewish population dataset26. In total, the number of SNPs used in ancestry estimation was 39,302 for samples genotyped on the NeuroBooster array and 24,404 for samples genotyped on the NeuroChip array. Ancestry estimates were carried out using a uniform protocol across all samples. A panel composed of 4008 samples from 1000 Genomes Project and the Gene Expression Omnibus (GEO) database were used (www.ncbi.nlm.nih.gov/geo (accession no. GSE23636) to define ancestry reference populations. The reference panel was then reduced to exclude palindromic SNPs (AT or TA or GC or CG). SNPs in the reference panel were further filtered to exclude variants with minor allele frequency (MAF) < 0.05, genotyping call rate < 0.99, and HWE P < 1E-4. Variants overlapping between the reference panel SNP set and the samples of interest were then extracted. Any missing genotypes were imputed using the mean of that particular variant in the reference panel.

The reference panel samples were split into an 80/20 train/test set and then principal components (PCs) were fit to and transformed via UMAP to represent global genetic population substructure and stochastic variation. A classifier was then trained on these UMAP transformations of the PCs (linear support vector). Based on the test data from the reference panel and at 5-fold cross validation, ancestries were predicted consistently with balanced accuracies greater than 0.95. These classifier models were then applied to GP2 and IPDGC data to generate ancestry estimates for all samples in the dataset. Samples predicted to have African or African admixed ancestry were then included in this meta-analysis. African admixed and African labels were combined into a single category and ADMIXTURE (v1.3.0; https://dalexander.github.io/admixture/binaries/admixture_linux-1.3.0.tar.gz) was run using the –supervised functionality to further divide these two categories where African is assigned if African admixture is >=90%, and African admixed is assigned if African admixture is <90%. For details of the cloud-based and scalable genotype calling, quality control and ancestry estimation pipeline, please see the GenoTools github repository (https://github.com/GP2code/GenoTools). Results from the respective GWA studies were annotated using ANNOVAR v2020-06-08 33. Samples clustering within African and African admixed ancestries were represented with 1000 Genome populations for visualization purposes (Supplementary Figures 2 and 3).

For the 23andMe dataset, an ancestry composition algorithm was applied by comparing 23andMe genome-wide content with an in-house customer reference panel composed of 14,393 individuals. Prior to this step, related individuals as well as individuals whose genetic ancestry did not match up with their survey answers were removed. A total of 45 ancestry composition populations were generated by performing principal component analysis using publicly available data from the African Genetics Project, the NIH-funded genetic health resource for African Americans, the Human Genome Diversity Project, HapMap, and the 1000 Genomes project, among others.

#### 23andMe data generation and processing

*23andMe genotyping*

Participants were genotyped on one of five genotyping platforms. The v1 and v2 platforms were based on the Illumina HumanHap550+ BeadChip, including about 25,000 custom variants selected by 23andMe, with a total of about 560,000 SNPs. The v3 platform was based on the Illumina OmniExpress+ BeadChip, with custom content to improve the overlap with our v2 array, with a total of about 950,000 variants. The v4 platform was a fully customized array, including a lower redundancy subset of v2 and v3 variants with additional coverage of lower-frequency coding variation, and about 570,000 variants. The v5 platform, in current use, is an Illumina Infinium Global Screening Array (~640,000 variants) supplemented with ~50,000 variants of custom content. This array was specifically designed to better capture global worldwide genetic diversity and to help standardize the platform for genetic research. Samples that failed to reach 98.5% call rate were re-genotyped. Participants whose analyses failed repeatedly were re-contacted by 23andMe customer service to provide additional samples. A total of 1,522,458 variants were genotyped across the five genotyping platforms.

*23andMe ancestry classifier*

A detailed description of 23andMe ancestry classifier can be found here (https://www.23andme.com/ancestry-composition-guide/). Briefly, the 23andMe ancestry classifier algorithm determines participant ancestries through an analysis of local ancestry30. It first partitions phased genotyped data into short windows of about 300 SNPs. Within each window, we use a support vector machine (SVM) to classify individual haplotypes into one of 45 worldwide reference populations. The SVM classifications are then fed into a hidden Markov model (HMM) that accounts for switch errors and incorrect assignments, and gives probabilities for each reference population in each window. Finally, we used simulated admixed individuals to recalibrate the HMM probabilities so that the reported assignments are consistent with the simulated admixture proportions. Each of the 45 reference populations were assigned to one of six higher-level populations: African American, East Asian, European, Latin American, and South Asian. The percentage of an individual’s ancestry derived from each of these six higher-level populations was calculated by summing HMM probabilities for all reference populations belonging to a given higher-level population.
African Americans and Latin Americans are admixed with broadly varying contributions from Europe, Africa, and the Americas. Therefore, no single threshold of genome-wide ancestry will be able to effectively discriminate African Americans and Latin Americans. However, the distributions of the length of segments of European, African and American ancestry are very different between African Americans and Latin Americans because of distinct admixture timing between the three ancestral populations in the two ethnic groups. Therefore, we trained a logistic classifier that takes one customer's length histogram of segments of African, European and American ancestry, and predicts whether the customer is likely African American or Latin American.

*23andMe phasing*

We built a high-quality phasing panel by selecting 200K African American participants with high quality genotyping (missingness < 5%). Variant QC statistics were computed independently for each genotyping platform, and the following criteria were used to select high quality variants:

- SNPs only (indels are excluded),
- Minor allele frequency (MAF) >= 0.1%,
- Missingness per variant < 5%,
- Correlation with sequence data > 0.9,
- MAF consistent with gnomAD (https://gnomad.broadinstitute.org/) (|Z|<100),
- Male heterozygosity tests for chromosome X non-PAR region.

All participant genotyping data were phased with SHAPEIT4 (https://odelaneau.github.io/shapeit4/) using genotyping platform specific phasing panels.

*23andMe imputation panels*

Imputation was performed using two independent reference panels: the publicly available Human Reference Consortium (HRC) panel and the 23andMe reference panel, which was built by 23andMe using internal and external cohorts.

HRC reference panel: The publicly available HRC data were downloaded from the European Genome-Phenome Archive at the European Bioinformatics Institute (accession EGAD00001002729). The HRC data includes 27,165 samples. Variants were liftovered to hg38 and excluded if their new positions were on a different chromosome. Variants were then re-phased using SHAPEIT4. Finally, singletons were excluded. The final HRC reference panel included 27,165 samples and 39,057,040 SNPs (no indels).

23andMe reference panel: We selected 12,217 samples from multiple internal and external WGS datasets. The cohort composition is included in **Supplementary Table 7.**

*23andMe imputation*

Phased genotyping data were used to impute 85M variants from the overall imputation panel for all participants using Beagle 531 Imputation was performed separately for the three sets of variants (HRC only, 23andMe only, and intersect), using three distinct sets of reference samples. Variants found in HRC only were imputed using the HRC reference samples. Variants found in 23andMe only were imputed using the 23andMe reference samples. Variants found in both HRC and 23andMe were imputed using all samples from both HRC and 23andMe. Imputation was performed independently for each 23andMe genotyping platform.

*Removing relatives in the 23andMe dataset*

A maximal set of unrelated individuals is chosen for each GWAS analysis using a segmental identity-by-descent (IBD) estimation algorithm32. Participants are defined as related if they shared more than 700 cM IBD, including regions where the two individuals share either one or both genomic segments IBD. This level of relatedness (roughly 20% of the genome) corresponds approximately to the minimal expected sharing between first cousins in an outbred population. When selecting individuals for case/control phenotype analyses, the selection process is designed to maximize case sample size by preferentially retaining cases over controls. Specifically, if both an individual case and an individual control are found to be related, then the case is retained in the analysis.

*23andMe GWAS*
We computed association test results for the genotyped and the imputed SNPs using logistic regression assuming additive allelic effects. For tests using imputed data, we used the imputed dosages rather than best-guess genotypes. As standard, we include covariates for age, sex, the top five principal components to account for residual population structure, and indicators for genotype platforms to account for genotype batch effects. The association test P value we report is computed using a likelihood ratio test. Results for the X chromosome are computed similarly, with male genotypes coded as if they were homozygous diploid for the observed allele.

*23andMe GWAS QC*
For QC of genotyped GWAS results, we flagged SNPs that were only genotyped on our “v1” and/or “v2” platforms due to small sample size, and SNPs on chrM or chrY because many of these are not currently called reliably. Using trio data, we flagged SNPs that failed a test for parent-offspring transmission; specifically, we regressed the child’s allele count against the mean parental allele count and flagged SNPs with fitted $\beta<0.6$ and P<10−20 for a test of $\beta<1$. We flagged SNPs with a call rate of <90%. We also tested genotyped SNPs for genotype date effects, and flagged SNPs with P<10−50 by ANOVA of SNP genotypes against a factor dividing genotyping date into 20 roughly equal-sized buckets. We flagged SNPs with large sex effect (ANOVA of SNP genotypes, r2>0.1). Finally, we flag SNPs with probes matching multiple genomic positions in the reference genome (‘self chain’).
For imputed GWAS results, we flagged SNPs with an imputation r2<0.5, as well as SNPs that had strong evidence of a platform batch effect. The batch effect test is an F test from an ANOVA of the SNP dosages against a factor representing v4 or v5 platform; we flagged results with P<10−50.
Across all results, we flag SNPs that have an available sample size of less than 20% of the total GWAS sample size. We also flag logistic regression results that did not converge due to complete separation, identified by abs(effect)>10 or stderr>10 on the log odds scale.

#### Short-read Whole Genome Sequencing

For Short-Read WGS, Psomagen quantified the starting genomic DNA material by fluorescence-based quantification method using Picogreen assay (cat# P7589, ThermoFisher) on VictorX2 multilabel plate reader (Perkin Elmer) and checked the DNA integrity using genomic DNA screen tape and reagents (cat. # 5067-5365/5067-5366, Agilent Technologies) on TapeStation 4200 (Agilent Technologies). A total of 0.5ug of gDNA was used as input material for Truseq PCR free library construction. The gDNA was fragmented to an insert size of 350 bp using LE220-plus Focused-ultrasonicator (Covaris). The generated double-stranded DNA with 3’or 5’overhangs was purified to get proper size range with sample purification beads. The size selected fragments were validated with D5000 ScreenTape (cat.# 5067-5588, Agilent Technologies) and D5000 Reagents (cat.# 5067-5589, Agilent Technologies) and then converted to blunt ends using end repair mix 2 from TruSeq DNA PCR-Free High Throughput Library Prep Kit (96 samples) (cat. # 20015963, illumina). One adenine (A) nucleotide was added to the 3’ ends of the blunt fragments to prevent them from ligating to each other during adapter ligation reaction. The adapter from IDT for Illumina – TruSeq DNA UD Indexes v2 (96 Indexes, 96 Samples) (cat.# 20040870 illumina) kit having one corresponding thymine (T) nucleotide on the 3' end was ligated to A-tailed DNA fragments. The adapter ligated fragments were then purified with sample purification beads to remove excessive reagents. The final purified library was validated with KAPA Library Quantification Kits (Cat.# 07960298001, Roche) on Light cycler 480 (Roche). The validated library was then normalized to 5nM and loaded onto Novaseq6000 after diluting to desired loading concentration (1.6nM).

### Global Parkinson's Genetics Program (GP2) Banner Authors

**Members: 207**
Version date: 16 March 2023

| Country | Name | Institution | Funders and Disclosures |
|---|---|---|---|
| Argentina | Emilia M Gatto | Sanatorio de la Trinidad Mitre - INEBA | Nothing to declare |
| | Marcelo Kauffman | Hospital JM Ramos Mejia | Nothing to declare |
| Armenia | Samson Khachatryan | Somnus Neurology Clinic | Nothing to declare |
| | Zaruhi Tavadyan | Somnus Neurology Clinic | Nothing to declare |
| Australia | Claire E Shepherd | Neuroscience Research Australia | The Sydney Brain Bank is located at and supported by Neuroscience Research Australia |
| | Julie Hunter | ANZAC Research Institute | Nothing to declare |
| | Kishore Kumar | Garvan Institute of Medical Research and Concord Repatriation General Hospital | Paul Ainsworth Family Foundation |
| | Melina Ellis | Concord Hospital | Nothing to declare |
| | Miguel E. Rentería | QIMR Berghofer Medical Research Institute | The Australian Parkinson's Genetics Study is supported by the Shake It Up Australia Foundation and The Michael J. Fox Foundation for Parkinson's Research |
| | Sulev Koks | Murdoch University | Nothing to declare |
| Austria | Alexander Zimprich | Medical University Vienna Austria | Nothing to declare |
| Brazil | Artur F. Schumacher-Schuh | Universidade Federal do Rio Grande do Sul / Hospital de Clínicas de Porto Alegre | Nothing to declare |
| | Carlos Rieder | Federal University of Health Sciences of Porto Alegre | Nothing to declare |

| | | | |
|---|---|---|---|
| | Vitor Tumas | University of São Paulo | Nothing to declare |
| Canada | Edward A. Fon | Montreal Neurological Institute | Nothing to declare |
| | Oury Monchi | Institut universitaire de gériatrie de Montréal | CIHR, Brain Canada, Parkinson Canada |
| | Ted Fon | McGill University | Nothing to declare |
| Chile | Benjamin Pizarro-Galleguillos | Faculty of Medicine Universidad de Chile | Nothing to declare |
| | Marcelo Miranda | Fundación Diagnosis | Nothing to declare |
| | Maria Leonor Bustamante | Fundación Diagnosis and Faculty of Medicine Universidad de Chile | Nothing to declare |
| | Patricio Olguin | Facultad de Medicina, Universidad de Chile | Nothing to declare |
| | Paula Saffie Awad | Universidade Federal do Rio Grande do Sul | Nothing to declare |
| | Pedro Chana | CETRAM, Universidad de Santiago de Chile | Nothing to declare |
| China | Beisha Tang | Central South University | Nothing to declare |
| | Huifang Shang | West China Hospital Sichuan University | Nothing to declare |
| | Jifeng Guo | Xiangya Hospital | Nothing to declare |
| | Piu Chan | Capital Medical University | Nothing to declare |
| | Wei Luo | Zhejiang University | Nothing to declare |
| | Xiaopu Zhou | The Hong Kong University of Science and Technology | Nothing to declare |
| Colombia | Gonzalo Arboleda | Universidad Nacional de Colombia | Nothing to declare |
| | Jorge Orozco | Fundación Valle del Lili | Nothing to declare |
| | Marlene Jimenez del Rio | University of Antioquia | Nothing to declare |
| Costa Rica | Alvaro Hernandez | University of Costa Rica | Nothing to declare |
| Egypt | Mohamed Salama | The American University in Cairo | The AUC/ ASRT/ DAAD |
| | Walaa A. Kamel | Beni-Suef University | Nothing to declare |

| | | | |
|---|---|---|---|
| Ethiopia | Yared Z. Zewde | Department of Neurology, College of Health Sciences, Addis Ababa University | Nothing to declare |
| France | Alexis Brice | Paris Brain Institute | Nothing to declare |
| | Jean-Christophe Corvol | Sorbonne Université | Nothing to declare |
| Germany | Brit Mollenhauer | University Medical Center Göttingen | Nothing to declare |
| | Christine Klein | University of Luebeck | CK serves as a medical Advisor to Centogene on genetic testing reports in the field of movement disorders, except Parkinson's disease, and is a member of the Scientific Advisory Board of Retromer Therapeutics |
| | Eva-Juliane Vollstedt | University of Luebeck | Nothing to declare |
| | Günter Höglinger | Department of Neurology, University Hospital, LMU Munich, Germany | Nothing to declare |
| | Prof. Franziska Hopfner | Department of Neurology, University Hospital, LMU Munich, Germany | Nothing to declare |
| | Harutyun Madoev | University of Luebeck | Nothing to declare |
| | Johanna Junker | University of Luebeck, Institute of Neurogenetics | Nothing to declare |
| | Katja Lohmann | University of Luebeck | Nothing to declare |
| | Lara M. Lange | University of Lübeck and University Medical Center Schleswig-Holstein | Nothing to declare |
| | Manu Sharma | University of Tubingen | Dr. Sharma is further funded by the Michael J Fox Foundation, USA Genetic Diversity in PD Program: GAP-India Grant ID: 009411 and 023430. Dr Sharma is supported by DFG (SH-599/16-1) |
| | Sergio Groppa | University of Mainz | Nothing to declare |
| | Thomas Gasser | University of Tübingen | Nothing to declare |

| | | | |
|---|---|---|---|
| | Zih-Hua Fang | The German Center for Neurodegenerative Diseases | Nothing to declare |
| Ghana | Albert Akpalu | University of Ghana Medical School | Nothing to declare |
| | Vida Obese | Queen Square Institute of Neurology | Nothing to declare |
| Greece | Georgia Xiromerisiou | University of Thessaly | Nothing to declare |
| | Georgios Hadjigorgiou | University of Thessaly | Nothing to declare |
| | Ioannis Dagklis | Aristotle University of Thessaloniki | Nothing to declare |
| | Ioannis Tarnanas | Ionian University | Nothing to declare |
| | Leonidas Stefanis | Biomedical research Foundation of the Academy of Athens | PPMI2 (funded by MJFF), ALAMEDA (H2020 grant), funding by HFRI |
| | Maria Stamelou | Diagnostic and Therapeutic Centre HYGEIA Hospital | Nothing to declare |
| | Efthimios Dardiotis | University of Thessaly | Nothing to declare |
| Honduras | Alex Medina | Hospital San Felipe | Nothing to declare |
| Hong Kong | Germaine Hiu-Fai Chan | Queen Elizabeth Hospital | Nothing to declare |
| | Nancy Ip | Hong Kong University of Science and Technology | Nothing to declare |
| | Nelson Yuk-Fai Cheung | Queen Elizabeth Hospital | Nothing to declare |
| | Phillip Chan | Hong Kong University of Science and Technology | Nothing to declare |
| India | Asha Kishore | Aster Medcity | Michael J Fox Foundation |
| | Divya KP | Sree Chitra Tirunal Institute for Medical Sciences and Technology | Nothing to declare |
| | Pramod Pal | National Institute of Mental Health & Neurosciences | Nothing to declare |
| | Prashanth Lingappa Kukkle | Manipal Hospital | Nothing to declare |
| | Roopa Rajan | All India Institute of Medical Sciences | Nothing to declare |

| | | | |
|---|---|---|---|
| | Rupam Borgohain | Nizam's Institute Of Medical Sciences | Nothing to declare |
| Italy | Andrea Quattrone | University "Magna Graecia" of Catanzaro | Nothing to declare |
| | Enza Maria Valente | University of Pavia, IRCCS Mondino Foundation | Nothing to declare |
| | Lucilla Parnetti | University of Perugia | Nothing to declare |
| | Micol Avenali | University of Pavia, IRCCS Mondino Foundation | Nothing to declare |
| | Tommaso Schirinzi | University of Rome Tor Vergata | Nothing to declare |
| Japan | Manabu Funayama | Juntendo University | Nothing to declare |
| | Nobutaka Hattori | Juntendo University faculty of medicine | Nothing to declare |
| | Tomotaka Shiraishi | Jikei University School of Medicine | Nothing to declare |
| Kazakhstan | Altynay Karimova | Institute of Neurology and Neurorehabilitation | Nothing to declare |
| | Rauan Kaiyrzhanov | University College London | Nothing to declare |
| | Gulnaz Kaishibayeva | Institute of neurology and neurorehabilitation named after Smagul Kaishibayev | Nothing to declare |
| Luxembourg | Rejko Krüger | University of Luxembourg | Nothing to declare |
| Malaysia | Ai Huey Tan | University of Malaya | Nothing to declare |
| | Azlina Ahmad-Annuar | University of Malaya | Nothing to declare |
| | Mohamed Ibrahim Norlinah | Universiti Kebangsaan Malaysia | Nothing to declare |
| | Nor Azian Abdul Murad | UKM Medical Molecular Biology Institute (UMBI) | Nothing to declare |
| | Norlinah Mohamed Ibrahim | Universiti Kebangsaan Malaysia Medical Centre | Nothing to declare |
| | Shahrul Azmin | Universiti Kebangsaan | Nothing to declare |

| | | | |
|---|---|---|---|
| | | Malaysia Medical Centre | |
| | Shen-Yang Lim | University of Malaya | Nothing to declare |
| | Wael Mohamed | International Islamic University Malaysia (IIUM) | Nothing to declare |
| | Yi Wen Tay | University of Malaya | Nothing to declare |
| Mexico | Daniel Martinez-Ramirez | Tecnologico de Monterrey | Nothing to declare |
| | Mayela Rodriguez-Violante | Instituto Nacional de Neurologia y Neurocirugia | Nothing to declare |
| | Paula Reyes-Pérez | Universidad Nacional Autónoma de México | Nothing to declare |
| Nepal | Rajeev Ojha | Tribhuvan University | Nothing to declare |
| New Zealand | Tim J Anderson | University of Otago, Christchurch, New Zealand;<br>New Zealand Brain Research Institute;<br>Neurology Department, Te Whatu Ora - Waitaha Canterbury, New Zealand | Health Research Council of New Zealand; Ministry of Business Innovation and Employment, New Zealand, Neurological Foundation of New Zealand |
| | Toni L Pitcher | University of Otago, Christchurch, New Zealand;<br>New Zealand Brain Research Institute | Health Research Council of New Zealand |
| Nigeria | Arinola Sanyaolu | University of Lagos | Nothing to declare |
| | Njideka Okubadejo | University of Lagos | Michael J Fox Foundation; Tertiary Education Trust Fund (TETFUND) National Research Fund |
| | Olaitan Okunoye | University College London | Nothing to declare |
| | Oluwadamilola Ojo | College of Medicine of the University of Lagos | Nothing to declare |
| Norway | Jan O. Aasly | Norwegian University of Science and Technology | Nothing to declare |
| | Lasse Pihlstrøm | Oslo University Hospital | Southeastern Regional Health Authority, Norway |

| | | | |
|---|---|---|---|
| | Manuela Tan | Oslo University Hospital | Southeastern Regional Health Authority, Norway<br>Michael J Fox Foundation |
| Pakistan | Shoaib Ur-Rehman | University of science and Technology Bannu | Nothing to declare |
| Peru | Miguel Inca-Martinez | Cleveland Clinic | Nothing to declare |
| | Mario Cornejo-Olivas | Neurogenetics Research Center, Instituto Nacional de Ciencias Neurologicas, Lima, Peru<br>Carrera de Medicina Humana, Universidad Cientifica del Sur, Lima, Peru. | MJFF and ASAP |
| Puerto Rico | Angel Vinuela | University of Puerto Rico | Nothing to declare |
| Russia | Elena Iakovenko | Research Center of Neurology | Nothing to declare |
| Saudi Arabia<br>Z | Bashayer Al Mubarak | King Faisal Specialist Hospital and Research Center | Nothing to declare |
| | Muhammad Umair | Medical Genomics Research Department, King Abdullah International Medical Research Center (KAIMRC), King Saud Bin Abdulaziz University for Health Sciences, Ministry of National Guard Health Affairs (MNGH), Riyadh, Saudi Arabia. | Nothing to declare |
| Singapore | Jia Nee Foo | Lee Kong Chian School of Medicine, Nanyang Technological University Singapore, Singapore<br>Genome Institute of Singapore, Agency | Singapore National Medical Research Council (MOH-000559) |

| | | | |
|---|---|---|---|
| | | for Science, Technology and Research (A*STAR), Singapore | |
| | Eng-King Tan | Department of Neurology, National Neuroscience Institute, Singapore Duke NUS Medical School, Singapore | National Medical Research Council Singapore (MOH-OFLCG-000207) |
| South Africa | Jonathan Carr | University of Stellenbosch | Nothing to declare |
| | Soraya Bardien | Division of Molecular Biology and Human Genetics, Stellenbosch University and The South African Medical Research Council/Stellenbosch University Genomics of Brain Disorders Research Unit | Funded by the National Research Foundation of South Africa [Grant Number 129249] |
| South Korea | Beomseok Jeon | Seoul National University Hospital | Nothing to declare |
| | Yun Joong Kim | Yongin Severance Hospital | Nothing to declare |
| Spain | Janet Hoenicka | Institut de Recerca Sant Joan de Deu | Fondo de Investigación Sanitaria, Instituto Salud Carlos III, Grant PI019/00126 |
| | Maria Teresa Periñan | Instituto de Biomedicina de Sevilla | Nothing to declare |
| | Pau Pastor | Unit of Neurodegenerative diseases, Department of Neurology, University Hospital Germans Trias i Pujol and Germans Trias i Pujol Research Institute (IGTP), Badalona, Barcelona, Spain | Nothing to declare |
| | Katrin Beyer | Germans Trias i Pujol Research Institute | Nothing to declare |

| | | | |
|---|---|---|---|
| | | (IGTP), Badalona, Barcelona, Spain | |
| | Ignacio Alvarez | Department of Neurology, Hospital Universitari Mutua de Terrassa, and Fundació per a la Recerca Biomèdica i Social Mútua de Terrassa, Terrassa, Spain. | Nothing to declare |
| Sudan | Sarah El-Sadig | Faculty of medicine university of Khartoum | Nothing to declare |
| Taiwan | Chin-Hsien Lin | National Taiwan University Hospital | Nothing to declare |
| | Hsiu-Chuan Wu | Chang Gung Memorial Hospital | Nothing to declare |
| | Pin-Jui Kung | Genome and systems biology degree program | Nothing to declare |
| | Ruey-Meei Wu | National Taiwan University Hospital | I have funding from 1. Minister of Science and Technology, Taiwan Government; 2. National Taiwan University, 3 Parkinson foundation, USA, 4. Michael J Fox Foundation. |
| | Serena Wu | Chang Gung University | Nothing to declare |
| | Yihru Wu | Chang Gung Memorial Hospital | Nothing to declare |
| Tunisia | Rim Amouri | National Institute Mongi Ben Hamida of Neurology | Nothing to declare |
| | Samia Ben Sassi | Mongi Ben Hmida National Institute of Neurology | Nothing to declare |
| Turkey | A. Nazlı Başak | Koç University | Kirac Foundation and Koc Univ. |
| | Gencer Genc | Şişli Etfal Training and Research Hospital | Nothing to declare |
| | Özgür Öztop Çakmak | Koç University | Nothing to declare |
| | Sibel Ertan | Koç University Medical School | Nothing to declare |

| | | | |
|---|---|---|---|
| United Kingdom | Alastair Noyce | Queen Mary University of London | Prof. Noyce reports grants from Parkinson's UK, Barts Charity, Cure Parkinson's, NIHR, Innovate UK, Virginia Keiley benefaction, Alchemab, Aligning Science Across Parkinson's and Michael J Fox Foundation. Consultancy and personal fees from Astra Zeneca, AbbVie, Profile, Roche, Biogen, UCB, Bial, Charco Neurotech, uMedeor and Britannia. |
| | Alejandro Martínez-Carrasco | University College London | Global Parkinson's Genetics Program |
| | Camille Carroll | University of Plymouth | C Carroll receives salary from University of Plymouth, University Hospitals Plymouth NHS Trust and National Institute of Health Research; she has received advisory, consulting, and/or lecture fees from AbbVie, Bial, Lundbeck, Global Kinetics, Britannia and Medscape, and research funding from Parkinson's UK, Edmond J Safra Foundation, National Institute of Health Research and Cure Parkinson's |
| | Claire Bale | Parkinson's UK | Nothing to declare |
| | Eleanor J. Stafford | University College London | Nothing to declare |
| | Henry Houlden | University College London | Nothing to declare |
| | Huw R Morris | University College London | Dr Morris is employed by UCL. In the last 12 months he reports paid consultancy from Roche and Amylyx ; lecture fees/honoraria - BMJ, Kyowa Kirin, Movement Disorders Society. Research Grants from Parkinson's UK, Cure Parkinson's Trust, PSP Association, CBD Solutions, Drake Foundation, Medical Research Council, Michael J Fox Foundation. Dr Morris is a co-applicant on a patent application related to C9ORF72 - Method for diagnosing a neurodegenerative disease (PCT/GB2012/052140) |

| | | | |
|---|---|---|---|
| | John Hardy | University College London | Nothing to declare |
| | Kin Ying Mok | University College London | Nothing to declare |
| | Mie Rizig | University College London | Nothing to declare |
| | Nicholas Wood | University College London | ASAP-CRN |
| | Nigel Williams | Cardiff University | Parkinson's UK |
| | Patrick Alfryn Lewis | Royal Veterinary College University of London | MJFF, UKRI (BBSRC, EPSRC), Parkinson’s UK, ASAP research network network |
| | Rimona Weil | University College London | Nothing to declare |
| | Simona Jasaitye | University College London | Nothing to declare |
| | Simon Stott | Cure Parkinson's | Employee of Cure Parkinson's |
| | Sumit Dey | Queen Mary University of London | Nothing to declare |
| USA | Alberto Espay | University of Cincinnati | Nothing to declare |
| | Alyssa O'Grady | The Michael J. Fox Foundation for Parkinson's Research | Nothing to declare |
| | Andrew B Singleton | National Institute on Aging | Michael J Fox Foundation for Parkinson's disease Research. Aligning Science Across Parkinson's Initiative |
| | Andrew K. Sobering | Augusta University / University of Georgia Medical Partnership | Nothing to declare |
| | Bernadette Siddiqi | The Michael J. Fox Foundation for Parkinson's Research | Nothing to declare |
| | Bradford Casey | The Michael J Fox Foundation for Parkinson's Research | Nothing to declare |
| | Brian Fiske | The Michael J. Fox Foundation for Parkinson's Research | Nothing to declare |
| | Cabell Jonas | Mid-Atlantic Permanente Medical Group / Kaiser Permanente Mid-Atlantic States | Nothing to declare |

| | | | |
|---|---|---|---|
| | Caroline B. Pantazis | National Institutes of Health | Nothing to declare |
| | Claire Wegel | Indiana University | Nothing to declare |
| | Cornelis Blauwendraat | National Institutes of Health | Nothing to declare |
| | Dan Vitale | National Institutes of Health | Nothing to declare |
| | Deborah Hall | Rush University | Nothing to declare |
| | Dena Hernandez | National Institutes of Health | Nothing to declare |
| | Ejaz Shiamim | Kaiser Permanente | Nothing to declare |
| | Ekemini Riley | Coalition for Aligning Science | Nothing to declare |
| | Faraz Faghri | National Institutes of Health | F.F.'s participation in this research was supported in part by the Intramural Research Program of the NIH, National Institute on Aging (NIA), National Institutes of Health, Department of Health and Human Services; project number ZO1 AG000535, as well as the National Institute of Neurological Disorders and Stroke. F.F.'s participation in this project was part of a competitive contract awarded to Data Tecnica International LLC by the National Institutes of Health to support open science research. |
| | Geidy E. Serrano | Banner Sun Health Research Institute | Banner Sun Health Research Institute Brain and Body Donation Program of Sun City, Arizona for the provision of human biological materials. The Brain and Body Donation Program has been supported by the National Institute of Neurological Disorders and Stroke (U24 NS072026 National Brain and Tissue Resource for Parkinson's Disease and Related Disorders), the National Institute on Aging (P30 AG19610 and P30AG072980, Arizona Alzheimer's Disease Center), the Arizona Department of Health Services (contract 211002, Arizona Alzheimer's Research Center), the Arizona Biomedical |

| | | | |
|---|---|---|---|
| | | | Research Commission (contracts 4001, 0011, 05-901 and 1001 to the Arizona Parkinson's Disease Consortium) and the Michael J. Fox Foundation for Parkinson's Research .” |
| | Hampton Leonard | National Institute on Aging/National Institutes of Health | H.L.L is supported by a competitive contract awarded to Data Tecnica International LLC by the National Institutes of Health to support open science research |
| | Hirotaka Iwaki | Data Tecnica International | Nothing to declare |
| | Honglei Chen | Michigan State University | NIH/DoD/Parkinson Foundation/MSU Foundation/Gibby vs. Parky Foundation - No COI to disclose |
| | Ignacio F. Mata | Cleveland Clinic | Funding from MJFF and NIH |
| | Ignacio Juan Keller Sarmiento | Northwestern University | Nothing to declare |
| | Jared Williamson | Kaiser Permanente | Nothing to declare |
| | Jonggeol Jeff Kim | National Institutes of Health | Nothing to declare |
| | Joseph Jankovic | Baylor College of Medicine | Nothing to declare |
| | Joshua Shulman | Baylor College of Medicine / Texas Children's Hospital | Collection of samples and data at Baylor College of Medicine was supported by the Huffington Foundation. |
| | Justin C. Solle | The Michael J. Fox Foundation for Parkinson's Research | Nothing to declare |
| | Kaileigh Murphy | The Michael J. Fox Foundation for Parkinson's Research | Nothing to declare |
| | Karen Nuytemans | University of Miami Miller School of Medicine | This work has been supported by the American Parkinson Disease Association and the Margaret Q. Landenberger Research Foundation. |

| | | | |
|---|---|---|---|
| | Karl Kieburtz | Beth Israel Deaconess Medical Center | Nothing to declare |
| | Kenneth Marek | Institute for Neurodegenerative Disorders | Consultant for Michael J Fox Foundation, GE Healthcare, Roche, UCB, BIAL, Denali, Takeda, , Cerapsir, UCB, Biohaven, Neuron23, Aprinoia, Astellas, Calico, Genentech, Invicro |
| | Kristin S. Levine | Data Tecnica International | K.S.L. is supported by a competitive contract awarded to Data Tecnica International LLC by the National Institutes of Health to support open science research |
| | Lana M. Chahine | University of Pittsburgh | Dr. Chahine receives research support from the Michael J Fox Foundation, UPMC Competitive Medical Research Fund, National Institutes of Health, and University of Pittsburgh, is study site investigator for a study sponsored by Biogen, receives consulting fees from Grey Matter Technologies, receives royalties from Elsevier (for authorship), and receives royalties from Wolters Kluwel (for authorship). |
| | Laurel Screven | National Institute on Aging | Nothing to declare |
| | Lisa Shulman | University of Maryland | Nothing to declare |
| | Luca Marsili | University of Cincinnati | Nothing to declare |
| | Maggie Kuhl | The Michael J. Fox Foundation for Parkinson's Research | Nothing to declare |

| | | | |
|---|---|---|---|
| | Marissa Dean | University of Alabama at Birmingham | Dr. Dean is an investigator in studies funded by Abbvie, Inc., Hoffmann-La Roche, CHDI Foundation, Inc., Annexon, Inc., Retrophin, Inc, Neurocrine Biosciences, UniQure Biopharma B.V., Praxis Precision Medicines, Neuraly, Inc., Michael J. Fox Foundation for Parkinson's Research, and US Army Medical Research and Materiel Command (grant#W81XWH-18-1-0508). In addition, Dr. Dean receives support through the Huntington's Disease Society of American Centers of Excellence program. |
| | Mary B Makarious | National Institutes of Health | Nothing to declare |
| | Mathew Koretsky | National Institutes of Health | Nothing to declare |
| | Megan J Puckelwartz | Northwestern University | Nothing to declare |
| | Mike A. Nalls | National Institutes of Health | M.A.N.'s participation in this project was part of a competitive contract awarded to Data Tecnica International LLC by the National Institutes of Health to support open science research. M.A.N. also currently serves as an advisor for Clover Therapeutics and Neuron23 Inc. |
| | Naomi Louie | The Michael J. Fox Foundation for Parkinson's Research | Nothing to declare |
| | Niccolò Emanuele Mencacci | Northwestern University | Nothing to declare |

| | | | |
|---|---|---|---|
| | Roy Alcalay | Columbia University | Dr. Alcalay is funded by the Michael J. Fox Foundation and the Parkinson's Foundation. He received consultation fees from Avrobio, Caraway, GSK, Merck, Sanofi, Ono Therapeutics and Takeda |
| | Sara Bandres-Ciga | National Institutes of Health | Nothing to declare |
| | Sohini Chowdhury | The Michael J. Fox Foundation for Parkinson's Research | Nothing to declare |
| | Sonya Dumanis | Aligning Science Across Parkinson's | Nothing to declare |
| | Steven Lubbe | Northwestern University | Nothing to declare |
| | Tao Xie | University of Chicago | Nothing to declare |
| | Tatiana Foroud | Indiana University School of Medicine | Michael J. Fox Foundation |
| | Thomas Beach | Sun Health Research Institution | Banner Sun Health Research Institute Brain and Body Donation Program of Sun City, Arizona for the provision of human biological materials. The Brain and Body Donation Program has been supported by the National Institute of Neurological Disorders and Stroke (U24 NS072026 National Brain and Tissue Resource for Parkinson's Disease and Related Disorders), the National Institute on Aging (P30 AG19610 and P30AG072980, Arizona Alzheimer's Disease Center), the Arizona Department of Health Services (contract 211002, Arizona Alzheimer's Research Center), the Arizona Biomedical Research Commission (contracts 4001, 0011, 05-901 and 1001 to the Arizona Parkinson's Disease Consortium) and the Michael J. Fox Foundation for Parkinson's Research ." |
| | Todd Sherer | The Michael J Fox Foundation for Parkinson's Research | Nothing to declare |

| | | | |
|---|---|---|---|
| | Yared Z. Zewde | University of California San Francisco | Nothing to declare |
| | Yeajin Song | National Institutes of Health | Nothing to declare |
| Vietnam | Duan Nguyen | Hue University | Nothing to declare |
| | Toan Nguyen | Hue University | Nothing to declare |
| Zambia | Masharip Atadzhanov | University of Zambia | Nothing to declare |

### Nigeria Parkinson Disease Research Network

| Surname | First name | Degree (preferred one specified) | Institutional affiliation (Name, City, State, Country) | ORCID |
|---|---|---|---|---|
| Agulanna | Uchechi | MBBS | Lagos University Teaching Hospital, Idi Araba, Lagos State, Nigeria | 0000-0002-0145-7833 |
| Akinyemi | Rufus | PhD | Neuroscience and Ageing Research Unit, Institute for Advanced Medical Research and Training, College of Medicine, University of Ibadan, Ibadan, Oyo State, Nigeria | 0000-0001-5286-428X |
| Ali | Mohammed | MBBS | Federal Teaching Hospital Gombe, Gombe State, Nigeria | 0000-0002-5596-9877 |
| Ani-Osheku | Ifeyinwa | FMCP | Asokoro District Hospital, Asokoro, Abuja, Nigeria | 0000-0002-1387-9806 |
| Arigbodi | Ohwotemu | MBBS | Delta State University, Abraka, Delta State, Nigeria | 0000-0001-9811-410X |
| Bello | Abiodun | FWACP | University of Ilorin Teaching Hospital, Ilorin, Kwara State, Nigeria | 0000-0002-2078-6739 |
| Erameh | Cyril | MBBS | Irrua Specialist Teaching Hospital, Irrua, Edo State, Nigeria | 0000-0001-7783-2495 |
| Farombi | Temitope | MBBS | University College Hospital, Ibadan, Oyo State, Nigeria | 0000-0002-9109-7164 |
| Fawale | Michael | MSc | Obafemi Awolowo University, Ile-Ife, Osun State, Nigeria | 0000-0003-3205-7514 |
| Imarhiagbe | Frank | MBCHB | University of Benin, Benin City, Edo State, Nigeria | 0000-0002-8518-2186 |
| Iwuozo | Emmanuel | FMCP | Benue State University, Makurdi, Benue State, Nigeria | 0000.0003-2378-3872 |
| Komolafe | Morenikeji | MBBS | Obafemi Awolowo University, Ile- Ife, Osun State, Nigeria. | 0000-0002-0592-5120 |
| Nwani | Paul | MBBS | Nnamdi Azikiwe University Teaching Hospital, Nnewi, Anambra State, Nigeria | 0000-0002-1665-2738 |
| Nwazor | Ernest | FMCP | Rivers State University Teaching Hospital, Port Harcourt, Rivers State, Nigeria | 0000-0001-7950-2449 |
| Nyandaiti | Yakub | MBBS | University of Maiduguri Teaching Hospital, Maiduguri, Borno State, Nigeria | 0000-0002-3922-6344 |
| Obiabo | Yahaya | MBCHB | Federal University of Health Sciences, Otukpo, Benue State, Nigeria | 0000-0003-4563-8908 |
| Odeniyi | Olanike | MBBS | General Hospital, Lagos Island, Lagos State, Nigeria | 0000-0002-8778-2230 |
| Odiase | Francis | MBBS | University of Benin, Benin City, Edo State, Nigeria | 0000-0001-9788-6482 |
| Ojini | Francis | MSc | University of Lagos, Lagos, Lagos State, Nigeria | 0000-0003-4108-4995 |
| Onwuegbuzie | Gerald | MBBS | University of Abuja, Abuja, Federal Capital Territory, Nigeria | 0000-0002-5203-1074 |
| Osaigbovo | Godwin | MBBS | Jos University Teaching Hospital, Jos, Plateau State, Nigeria | 0000 0001 5572 0548 |
| Osemwegie | Nosakhare | MBBS | University of Port Harcourt, Port Harcourt, Rivers State, Nigeria | 0000-0002-8725-2256 |
| Oshinaike | Olajumoke | FWACP | Lagos State University College of Medicine, Ikeja, Lagos State, Nigeria | 0000-0002-6290-6866 |
| Otubogun | Folajimi | MBCHB | Federal Medical Center, Ebute Metta, Lagos State, Nigeria | 0000-0001-9753-4532 |
| Oyakhire | Shyngle | MBBS | National Hospital, Abuja, Federal Capital Territory, Nigeria | 0000-0002-8442-9896 |
| Ozomma | Simon | FMCP | University of Calabar Teaching Hospital, Calabar, Cross River State, Nigeria | 0009-0001-6921-3544 |
| Samuel | Sarah | MBBS | University of Maiduguri Teaching Hospital, Maiduguri, Borno State, Nigeria. | 0000-0001-6178-8282 |
| Taiwo | Funmilola | MBCHB | University College Hospital, Ibadan, Oyo State, Nigeria | 0000-0002-5797-0943 |
| Wahab | Kolawole | MD | University of Ilorin, Ilorin, Kwara State, Nigeria | 0000-0002-2914-1953 |
| Zubair | Yusuf | MSc | National Hospital, Abuja, Federal Capital Territory, Nigeria | 0000-0002-3017-7961 |

**International Parkinson Disease Genomics Consortium Africa members by country**

**Cameroon**

1. Daniel Gams Massi, MD. Douala General Hospital, University of Buea, Cameroon.
2. Eric Gueumekane Bila lamou, MD. University of Douala, Douala, Cameroon.
3. Leonard Njamnshi Nfor, MD. Yaounde Central Hospital, Yaoundé, Cameroon.
4. Mélanie Annick Magnerou, MD. Douala Gynaeco-Obstetric and Paediatric Hospital, Douala, Cameroon.
5. Yannick Fogang Fogoum, MD. Bafoussam Regional Hospital, Bafoussam, Cameroon.

**Egypt**

1. Ali shalash, MD. Neurology Department, Ain Shams University, Cairo, Egypt.
2. Hassan El-Fawal, PhD. Institute of Global Health and Human Ecology, the American University in Cairo, Egypt.
3. Eman Khedr, MD. Neurology Department, Assiut University, Assiut, Egypt.
4. Gharib Fawi, MD. Neurology Department, Sohag University, Sohag, Egypt.
5. Mohamed A. Eltantawi, MD. Neurology Department, Delta University for Science and Technology, Belkas, Dakahlia Governorate, Egypt.
6. Mohamed Salama, PhD. Institute of Global Health and Human Ecology, the American University in Cairo, Egypt.
7. Shaimaa El-Jaafary, MD. Neurology Department, Cairo University, Cairo, Egypt.
8. Sharifa Hamed, MD. Neurology Department, Assiut University, Assiut, Egypt.

**Ethiopia**

1. Abenet Tafesse Mengesha, MD. Department of Neurology, School of Medicine College of Health Sciences, Addis Ababa University, Addis Ababa, Ethiopia.
2. Biniyam Alemayehu Ayele, MD. Department of Neurology, School of Medicine College of Health Sciences, Addis Ababa University, Addis Ababa, Ethiopia.
3. Dereje Melka Oda, MD. Department of Neurology, School of Medicine College of Health Sciences, Addis Ababa University, Addis Ababa, Ethiopia.
4. Yared Zenebe Zewde, MD. Department of Neurology, School of Medicine College of Health Sciences, Addis Ababa University, Addis Ababa, Ethiopia.
5. Yohanesse Debebe Gelan, MD. Department of Neurology, School of Medicine College of Health Sciences, Addis Ababa University, Addis Ababa, Ethiopia.

**Ghana**

1. Albert Akpalu,FRCP. Department of Medicine, Division of Neurology, University of Ghana Medical School, Accra, Ghana.
2. Augustina Charway-Felli, PhD. Department of Medicine, Division of Neurology, 37 Military Hospital, Accra, Ghana.
3. Fred Stephen Sarfo, PhD. Department of Medicine, Division of Neurology, Kwame Nkrumah University of Science & Technology, Kumasi, Ghana.
4. Patrick Adjei, PhD. Department of Medicine, Division of Neurology, University of Ghana Medical School, Accra, Ghana.
5. Vida Obese, MD. Department of Medicine, Division of Neurology, Komfo Anokye Teaching Hospital, Kumasi, Ghana.

**Mali**

1. Abdoulaye Bocoum, MD. University of Sciences, Technics and Technologies of Bamako, Mali.
2. Abdou Koita, MD. University of Sciences, Technics and Technologies of Bamako, Mali.
3. Cheick Oumar Guinto, MD. University of Sciences, Technics and Technologies of Bamako, Mali; Department of Neurology, Teaching Hospital of Point "G", Bamako, Mali.
4. Toumany Coulibaly, MD. Department of Neurology, Teaching Hospital of Point "G", Bamako, Mali.
5. Youssoufa Maiga, MD. University of Sciences, Technics and Technologies of Bamako, Mali; Department of Neurology, Teaching Hospital of Gabriel Touré, Bamako, Mali.
6. Zaynab Kone, MD. Department of Neurology, Teaching Hospital of Point "G", Bamako, Mali.

**Nigeria**

1. Abiodun Bello, FWACP. University of Ilorin Teaching Hospital, Ilorin, Nigeria.
2. Agabi Osigwe, FMCP. consultant Neurologist with the Lagos University Teaching Hospital, Nigeria.
3. Akintunde A. Adebowale, FWACP. Obafemi Awolowo University, Ile-Ife, Osun State. Nigeria
4. Akpekpe John, FWACP. Federal Medical Center, Asaba, Delta State, Nigeria.
5. Alagoma Iyagba, MBBS. University of Port Harcourt and University of Port Harcourt Teaching Hospital, port Harcourt, Rivers State. Nigeria.
6. Ali Mohammed Wulgo, FWACP. Federal Teaching Hospital, Gombe, Gombe State. Nigeria
7. Ani-Osheku Ifeyinwa, FMCP. Asokoro District Hospital, Asokoro, Federal Capital Territory, Abuja. Nigeria.
8. Babawale Arabambi, FWACP. Lagos State University Teaching Hospital. Nigeria.
9. Charles Achoru, FWACP. University Teaching Hospital, Jos, Plateau State. Nigeria.
10. Christian Ejikeme Agu Alex Ekwueme, Federal Teaching Hospital, Abakaliki, Ebonyi State. Nigeria.
11. Cyril Erameh, FMCP. Irrua Specialist Hospital, Irrua, Edo State. Nigeria.
12. Emmanuel Uzodinma Iwuozo, FMCP. Medicine department, College of Health Sciences Benue State University, Benue State. Nigeria.
13. Ernest. O. Nwazor. FWACP Federal Medical Center, Owerri, Imo State & College of Medical Sciences, Madonna University, Elele, Rivers State. Nigeria.
14. Fawale Michael Bimbola, FMCP. Obafemi Awolowo University and Obafemi Awolowo University Teaching Hospitals Complex, Ile-Ife, Osun State. Nigeria.
15. Folajimi Otubogun, FWACP. University of Medical Sciences Teaching Hospitals Complex, Akure, Ondo State. Nigeria.
16. Francis Odiase, FMCP. University of Benin, University of Benin Teaching Hospital, Benin city Edo State, Nigeria.
17. Franklin Dike, FWACP. University of Uyo Teaching Hospital, Uyo, Akwa Ibom State. Nigeria.
18. Ishola Ismail O. Ph.D. Department of Pharmacology, Lagos State University and Lagos State University Teaching Hospital, Lagos State. Nigeria.
19. Kolawole Wahab, FMCP. University of Ilorin. Ilorin.Nigeria.
20. Kehinde Johnson Abiodun, FMCP. Consultant Neurologist, Federal Medical Centre, Abuja, FCT, Nigeria
21. Morenikeji Komolafe, FWACP. Department of Medicine, Faculty of Clinical Sciences, College of Medicine, Obafemi Awolowo University, Ile-Ife, Osun State, Nigeria.
22. Njideka Okubadejo, FMCP. Neurology Unit, Department of Medicine, Faculty of Clinical Sciences, College of Medicine, University of Lagos, Lagos State, Nigeria

23. Obiabo Yahaya Olugbo, FMCP. Neurology Unit, Department of Internal Medicine, Delta State, University Teaching Hospital, Oghara. Nigeria
24. Ohwotemu Arigbodi, FMCP. Delta state University Teaching hospital, Oghara. Nigeria.
25. Olajumoke Oshinaike, FWACP. Lagos State University & Lagos State University Teaching Hospital, Lagos State. Nigeria.
26. Osemwegie Nosakhare, FMCP. University of Port Harcourt and University of Port Harcourt Teaching Hospital. Nigeria
27. Oluchi Ekenze, FWACP. University of Nigeria & University of Nigeria Teaching Hospital, Ituku-Ozalla, Enugu State. Nigeria
28. Oluwadamilola Ojo, FMCP. University of Lagos & Lagos University Teaching Hospital, Lagos State. Nigeria
29. Paul Agabi Osigwe, FMCP. Lagos University Teaching Hospital, Lagos State. Nigeria.
30. Paul O Nwani, FWACP. Neurology Unit, Dept. of Medicine, Faculty of Medicine, Nnamdi Azikiwe University, Nnewi, Anambra State, Nigeria.
31. Salisu Abdullahi Balarabe, FWACP. Usmanu Danfodiyo University Teaching Hospital Sokoto, Nigeria
32. Sani Abubakar Atta Ahmadu Bello, WACP. University and Ahmadu Bello University Teaching Hospital, Zaria, Kaduna State. Nigeria
33. Uduak E. Williams, FMCP. University of Calabar and University of Calabar Teaching Hospital, Calabar, Nigeria.
34. Oladunni V Abiodun, FMCP, General Hospital Isolo, Isolo, Lagos State. Nigeria
35. Yusuf Agboola Zubair, FWACP. National Hospital, Federal Capital Territory, Abuja.

**Senegal**

1. Maouly Fall, MD. Centre Hospitalier National de Pikine and Cheikh Anta Diop University, Senegal.
2. Alassane Mamadou Diop, MD. Centre Hospitalier National de Pikine, Senegal.
3. Ewodo Touna Hilaire Dominique, MD. Neurological Clinic Ibrahima P. Ndiaye, Fann Hospital University – Cheikh Anta Diop University of Dakar (UCAD), Dakar, Senegal.

**South Africa**

1. Andre Mochan, MD. University of the Witwatersrand, Neurosciences, Division of Neurology, Johannesburg, South Africa.
2. Girish Modi, PhD. University of the Witwatersrand, Neurosciences, Division of Neurology, Johannesburg, South Africa.
3. Saiesha Dindayal, FC Neurol (SA). University of the Witwatersrand, Neurosciences, Division of Neurology, Johannesburg, South Africa.

**Sudan**

1. Eman Ali Awadelkareem, MSc. Sudan University of Science and Technology, Sudan.
2. Inas Elsayed, MSc. Faculty of Pharmacy, University of Gezira, Sudan.
3. Maha Dahawi, MSc. Faculty of Medicine, University of Khartoum, Sudan.
4. Mosab Ali Awadelkareem, MSc. Faculty of Medical Laboratory Sciences, Al-Neelain University, Sudan.
5. Sarah Misbah, MRCP. Faculty of Medicine, University of Khartoum, Sudan.

**Tanzania**

1. Brighton Mushengezi, MD. Muhimbili National Hospital, Upanga Branch. Tanzania.
2. Henrika Kimambo, MD. Muhimbili National Hospital UPanga Branch. Tanzania.
3. Leonard Msango, MD. Muhimbili National Hospital, Mloganzile Branch. Tanzania.
4. Philip Adebayo, FWACP. Aga Khan Hospital and Aga Khan University. Tanzania.
5. Kigocha OKengo, MD. Muhimbili National Hospital UPanga Branch. Tanzania
6. Marieke Diekker, PhD, Kilimanjaro Christian Medical Center. Kilimanjaro. Tanzania.
7. Sarah URassa, MSc. Kilimanjaro Christian Medical Center. Kilimanjaro. Tanzania.

**Tunisia**

1. Riadh Gouider, MD. Department of Neurology, Clinical Investigation Center, Razi Hospital, Faculty of Medicine of Tunis, Tunis El Manar University, Tunisia.
2. Mouna Ben Djebara, MD. Department of Neurology, Clinical Investigation Center, Razi Hospital, Faculty of Medicine of Tunis, Tunis El Manar University, Tunisia.
3. Amina Gargouri, MD. Department of Neurology, Clinical Investigation Center, Razi Hospital, Faculty of Medicine of Tunis, Tunis El Manar University, Tunisia.
4. Imen Kacem, MD. Department of Neurology, Clinical Investigation Center, Razi Hospital, Faculty of Medicine of Tunis, Tunis El Manar University, Tunisia.
5. Amina Nasri, MD. Department of Neurology, Clinical Investigation Center, Razi Hospital, Faculty of Medicine of Tunis, Tunis El Manar University, Tunisia.
6. Saloua Mrabet, MD. Department of Neurology, Clinical Investigation Center, Razi Hospital, Faculty of Medicine of Tunis, Tunis El Manar University, Tunisia.
7. Ikram Sghaier, PhD in Biology (human genetics), Department of Neurology, Clinical Investigation Center, Razi Hospital, Tunis - Tunisia
8. Imen Mkada, PhD. Department of Neurology, Clinical Investigation Center, Razi Hospital, Tunis - Tunisia

**Zambia**

1. Masharip Atadzhanov, PhD. University of Zambia, School of Medicine, Department of Internal Medicine, Lusaka, Zambia
2. Lorraine Chishimba, MMED Adult neurology. University Teaching Hospital, Department of Internal Medicine, Lusaka, Zambia.

**UK**

1. Mie Rizig, PhD. Department of Neuromuscular Diseases, UCL Queen Square Institute of Neurology, Queen Square London WC1N 3BG, UK.
2. Fatumah Jama, BSc. Department of Neuromuscular Diseases, UCL Queen Square Institute of Neurology, Queen Square London WC1N 3BG, UK.
3. Olaitan Okunoye, MSc. Department of Movement Disorders, UCL Queen Square Institute of Neurology, Queen Square London WC1N 3BG, UK.
4. John Hardy, PhD. Department of Neurodegenerative Diseases, UCL Queen Square Institute of Neurology, Queen Square London WC1N 3BG, UK.
5. Henry Houlden, PhD. Department of Neuromuscular Diseases, UCL Queen Square Institute of Neurology, Queen Square London WC1N 3BG, UK.

**USA**

1. Andrew Singleton, PhD. Laboratory of Neurogenetics, National Institute on Aging, National Institutes of Health, Bethesda, MD 20892, USA.

2. Mike Nalls, PhD. Laboratory of Neurogenetics, National Institute on Aging, National Institutes of Health, Bethesda, MD 20892, USA; and Data Tecnica International, Glen Echo, MD, USA.

#### BLAAC PD Study Group Members

**Ejaz Shamim,** MD - Department of Neurology, Kaiser Permanente Mid-Atlantic States, Rockville, Maryland
Cabell Jonas, PhD
Jared Williamson

**Deborah A. Hall,** MD, PhD - Department of Neurology, Rush University Medical Center, Chicago, Illinois
Marc Rosenbaum, MS, CGC
Staci Davis, BS

**Marissa Dean, MD and David G Standaert, MD, PhD** - Department of Neurology, University of Alabama at Birmingham
Candace Cromer, BSc
Jenna Smith, RN
Joseph Richardson, RN
Lauren Ruffrage
Rebeka Sipma, MD

**Tao Xie, MD, PhD** - Department of Neurology, University of Chicago Medicine, Chicago, Illinois, USA
Manesh Padmanaban, MD
Natalie Warren, BSc
Tomas Mercado

**Elizabeth Disbrow, PhD** - Center for Brain Health, LSU Health Shreveport, Shreveport, LA 71130, USA; Department of Neurology, LSU Health Shreveport, Shreveport, LA 71130, USA
Brian Chauppeta
Fermine Thomas-Dean, PhD
Jamie Toms, MD
Katelyn Lofton, BSc

**Ashley Rawls, MD, MS** - Department of Neurology, University of Florida College of Medicine, Gainesville, FL-32611, USA
Kyle Rizer
Nieci Black, MPH

**The Michael J Fox Foundation for Parkinson's Research**
Justin Solle, MBA
Alyssa O’Grady, BSc
Todd Sherer, PhD
Brian Fiske, PhD
